# Air Pollution and Children’s Health Inequalities

**DOI:** 10.1101/2024.02.07.24302381

**Authors:** Milena Suarez Castillo, David Benatia, Christine Le Thi, Vianney Costemalle

## Abstract

This paper examines the differential impacts of early childhood exposure to air pollution on children’s health care use across parental income groups and vulnerability factors using French administrative data. Our quasi-experimental study reveals significant impacts on emergency admissions and respiratory medication in young children, attributed to air pollution shocks from thermal inversions. Using causal machine learning, we identify these health impacts as predominantly affecting 10% of infants, characterized by poor health indicators at birth and lower parental income. Our results suggest that intervention strategies focusing on vulnerability metrics may be more effective than those based solely on exposure levels.

*JEL Codes*: I14, I18, Q53, Q58

## 1 Introduction

Air quality and environmental inequalities are urgent health policy issues, especially when young children are at risk. The *average* impacts of air pollution on mortality, morbidity (Currie et al., 2014; Velasco and Jarosińska, 2022), and its links to childhood asthma are well-documented (Khreis et al., 2019). However, the picture is far from complete since air pollution does not affect all children equally. Knowledge about the differential effects of early childhood exposure to air pollution across varied socio-economic backgrounds remains scarce. This gap is notable as children across these backgrounds display differences in vulnerability, exposure to pollution, and accessibility to health care.

Figure 1a shows a marked reduction in the average yearly fine particulate matter (PM2.5) exposure for children in France from 2008 to 2017, using two distinct data sources, based on satellite measurements (ACAG) or on emission inventories (Ineris). Figure 1b reveals significant inequalities in exposure: children from both the upper and lower parental income deciles experience the highest pollution exposure within their birth cohorts.

**Figure 1:**
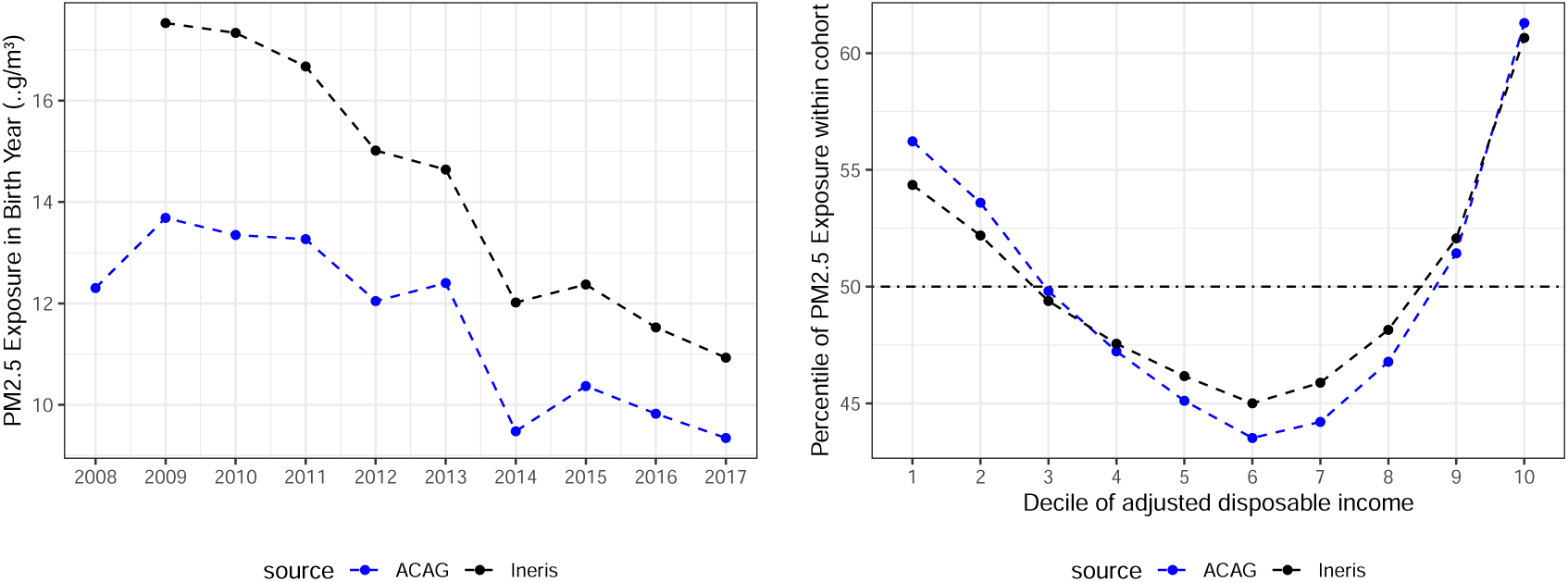
Average PM2.5 exposure by birth year and within decile of adjusted household income. *Source:* ACAG: Atmospheric Composition Analysis Group; Ineris: French National Institute for Industrial Environment and Risks; EDP-Santé

This paper studies the heterogeneous health consequences of temporary increases in exposure to PM2.5 for children using quasi-experimental evidence linking health outcomes to socio-demographics, vulnerability factors, and air pollution exposure. To investigate these inequalities, we leverage a large representative administrative data sample of about 340,000 children born in France between 2008 and 2017, which includes comprehensive information on their health care utilization and parental income, and use generic machine learning inference (Chernozhukov et al., 2023) to identify the groups that are particularly vulnerable to the health consequences of air pollution exposure.

Identifying the causal effects of air pollution exposure on children’s health care utilization is challenging due to three main endogeneity issues. First, pollution, as a by-product of economic activity, not only affects health but is also correlated with other factors such as work conditions and habits (Dehejia and Lleras-Muney, 2004; Heutel and Ruhm, 2016; Stevens et al., 2015). Second, household location choices are endogenous dynamic decisions that may be influenced by anticipated air pollution exposure and related amenities like the absence of major roads (Pan, 2023). Finally, measurement errors in individual pollution exposure, with precise daily tracking being impractical and concentration measures showing significant source-dependent variations, can introduce an attenuation bias.

To address these endogeneity issues, we use a quasi-experimental design that isolates a particular type of air pollution shock: local exposure to thermal inversions. This phenomenon arises when a layer of warm air traps cooler air beneath it, preventing the dispersal of pollutants and leading to elevated levels of PM2.5 in the surrounding area. Our empirical strategy can be described as a reduced-form instrumental variable (IV) analysis; or intention-to-treat (ITT) framework (Imbens, 2014); where we design a binary treatment from a plausibly exogenous variable, to implement a rich heterogeneity analysis following Chernozhukov et al. (2023). We categorize infants into “more exposed” (treated) and “less exposed” (control) groups depending on their exposure to an above-average number of thermal inversions in their first year, factoring in municipality-specific and year-specific averages. This group distinction offers credible exogenous variations in air pollution exposure, evidenced by its significant association with fine particulate matter concentrations and its lack of correlation with individual child characteristics. Our causal estimations hinge on the assumption that, after controlling for municipality and year fixed-effects as well as weather variables, changes in the number of thermal inversions within a municipality are unrelated to changes in infant health care use, except through their influence on air pollution.

Our analysis examines two key indicators of health care use linked to air pollution: emergency admissions, and medication dispensed for respiratory conditions. More specifically, we focus on emergency admissions and medications related to bronchiolitis and asthma.^1^ In 2017, medications for obstructive airway diseases accounted for EUR 1 billion in reimbursements from the French National Health Insurance, serving 8.6 million beneficiaries, 26.6% of whom were under 20 years old.^2^ France’s comprehensive universal health care system minimizes financial barriers, making health service usage a more accurate reflection of actual health care needs compared to many OECD countries (OECD, 2019). This universal system facilitates not only the analysis of drug prescriptions but also provides access to comprehensive health care utilization data across a representative population sample, contrasting with the more limited scope of data from specific insurance groups or Medicare (Klauber et al., 2023; Deryugina et al., 2019). This broad and representative coverage renders our dataset ideal for investigating income-based health disparities.

Our findings indicate that infants overexposed to air pollution in their first year have *on average* a higher propensity to utilize healthcare services for respiratory issues compared to their less exposed peers. After controlling for municipality and year fixed-effects and weather variables, the infants in the exposed birth cohorts experience an average of 11 additional days with thermal inversion in their first year (+30%). This corresponds to a short-term spike in PM2.5 pollution exposure over a few days and equates to a 0.1 to 0.2 *µg/m*^3^ increase in average *annual* exposure, or about 1 to 2%. Our results indicate that the exposed cohorts are more likely to experience at least one emergency admission for bronchiolitis or asthma during their first three years, with an increased probability of 0.5 p.p. (12% of the mean probability). These children also show a higher propensity for anti-asthma drug consumption before their first birthday, a probability that increases with the extent of air pollution exposure differential.

We extend this analysis by exploring various dimensions of heterogeneity, notably parental income and vulnerability at birth. Our results reveal that the impacts of air pollution are concentrated in about 10% of the infant population, which are mainly characterized by a combination of poor health indicators at birth. The most impacted infants (top 10%) exhibit a 2.5 p.p. increased likelihood of being prescribed anti-asthma medication (10% of their group-specific mean probability) and a 1.3 p.p. increased likelihood of at least one emergency bronchiolitis admission (27% of their group-specific mean probability) if exposed to the air pollution shock within their first year.

Our results also hint that air pollution is a leading cause of emergency admissions for bronchiolitis among the bottom and top deciles of parental income. Figure 2 reveals a negative gradient between the likelihood of emergency admissions for bronchiolitis and parental income, with the bottom and top deciles accounting for 14% and 6.7% of all admissions, respectively. However, for admissions attributed to air pollution, these percentages rise to 17.4% for the lowest income group and 11.1% for the highest, emphasizing the disproportionate impact of air pollution on admissions for both ends of the income spectrum.

**Figure 2:**
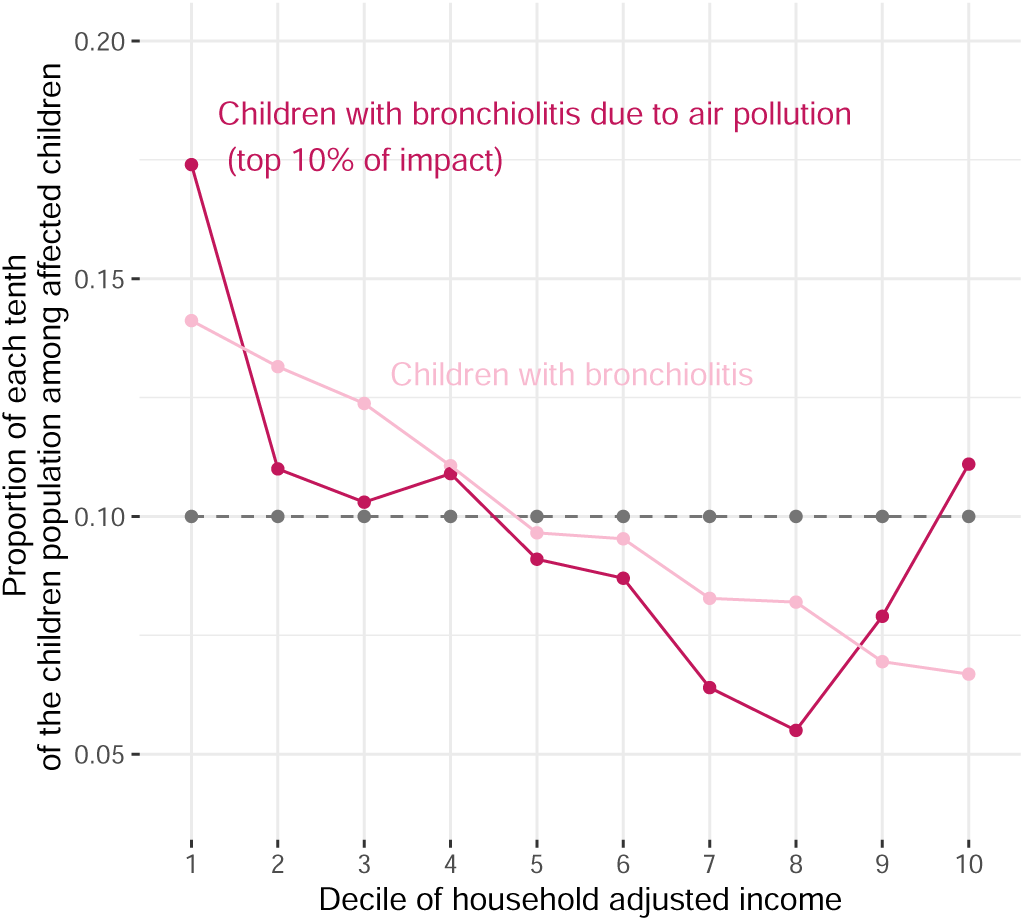
Distribution of children household adjusted income in children with an emergency admission for bronchiolitis (all causes) and in children with an emergency admission for bronchiolitis due to air pollution, as identified by our heterogeneity analysis.

Our findings underscore that temporary surges in air pollution exposure contribute to respiratory health complications in young children, with the most vulnerable being disproportionately affected. This highlights the urgent need for mitigation policies that extend beyond traditional exposure level considerations, calling for a tailored approach recognizing local characteristics of population and pollution sources. In our discussion, we employ the European Union (EU)’s proposed amendments to the Ambient Air Quality Directives both as a framework for discussion and to evaluate mitigation policies aimed at achieving the EU’s interim air pollution exposure targets for 2030.^3^ Our research suggests the need for more precise air quality monitoring, the development of harmonized indices that account for population-specific vulnerabilities, and targeted compensation schemes, particularly for lower-income groups. Crucially, our empirical analysis informs the development of more effective national air quality roadmaps, aligning with the EU’s mandate. It underscores that prioritizing regions with vulnerable populations, identified by simple metrics like the number of premature births, may more effectively mitigate the adverse health impacts of air pollution compared to approaches based solely on pollution exposure. This vulnerability-based targeting could emerge as a more effective strategy over exposure-based prioritization.

Our research engages with the literature exploring the heterogeneous health impacts on air pollution. Deryugina et al. (2019) find that the mortality effects of air pollution are concentrated in the most vulnerable 25% of the elderly population, by leveraging the generic machine learning approach (Chernozhukov et al., 2023), whereas Deryugina et al. (2021) examine the associated socio-economic and geographical determinants. We consider that infants and young children warrant particular attention in the study of health disparities stemming from air pollution due to the potential lifelong consequences (Isen, Rossin-Slater and Walker, 2017; Bharadwaj et al., 2017) and intergenerational transmissions of inequalities (Currie, 2009). In this line of research, and closely related to our work, Jans, Johansson and Nilsson (2018) document the impact of air quality on infant health by socioeconomic status, using thermal inversions to tackle endogenous residential sorting. Thermal inversions have been also used as an exogenous source of variations for air pollution in Arceo, Hanna and Oliva (2016) and Dechezleprêtre, Rivers and Stadler (2019), among others, to investigate related environmental questions.

The remainder of this paper is organized as follows. Section 2 presents some background facts about inequalities and air pollution in France. Section 3 describes the data used in our empirical study. Section 4 presents the empirical strategy. Section 5 collects the empirical results and related discussion. Section 6 discusses policy implications. Section 7 concludes.

## 2 Health Inequalities & Air Pollution in France

Children in dense urban areas are disporportionately exposed to fine particulate matter, highlighting the environmental inequalities faced by some communities. Figure 3 shows the high concentration of PM2.5 in these populous areas, including France’s major cities, as depicted in Figure A3. This phenomenon reflects broader socioeconomic disparities beyond the simple urban-rural divide. Nationally, children from both the lower and upper parental income deciles experience elevated air pollution levels, indicative of spatial sorting effects. The upper decile is more exposed due to their over-representation in the largest, often more polluted, French urban areas. Nonetheless, within these areas, children from poorer backgrounds face the greatest exposure to PM2.5, as shown in Figure A2, a consequence of their residence in the most polluted municipalities.

**Figure 3:**
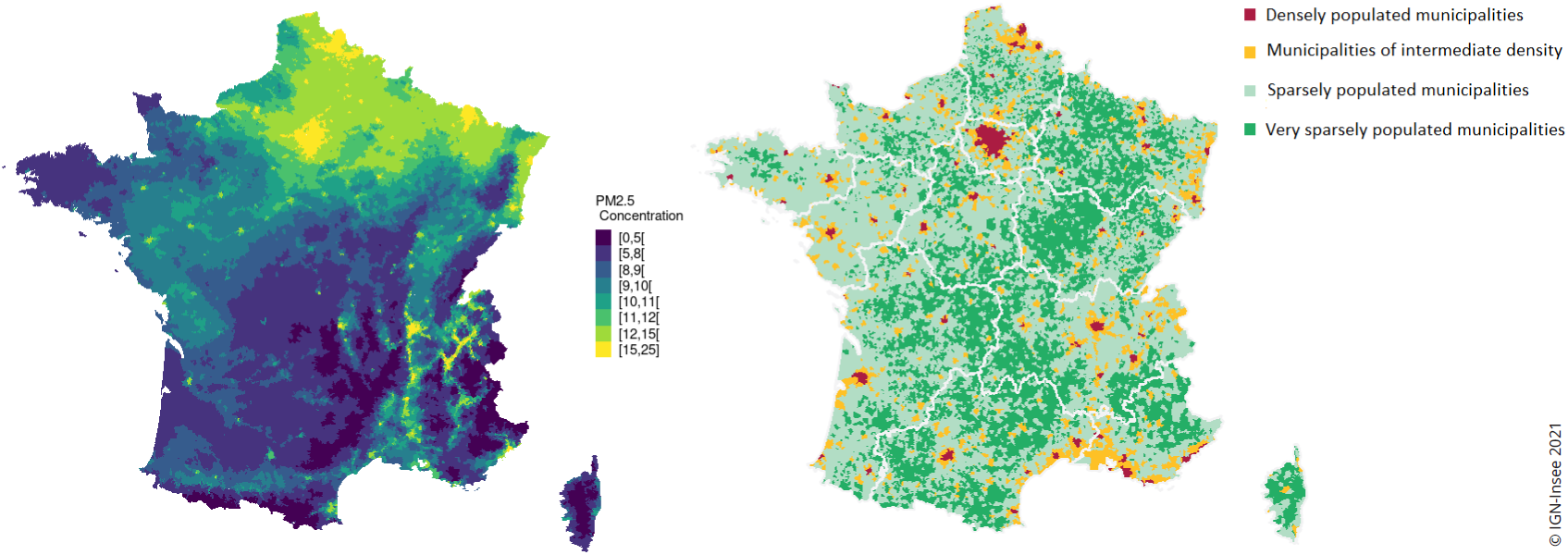
PM2.5 concentrations at the municipality level (*µ*g/m^3^) in 2010 (source: ACAG) and population densities in 2017 (source: Insee-IGN, 2021)

Recent U.S. research indicates that higher exposure to air pollution persists among specific racial groups and, to some degree, low-income populations (Currie, Voorheis and Walker, 2023), suggesting that health inequality due to air pollution is a global issue, not unique to France. Hsiang, Oliva and Walker (2019) observed that although cities with higher household incomes have increased levels of nitrogen dioxide (NO2), it is the disadvantaged groups within these cities who endure the most exposure. The study also reveals a U-shaped relationship between income and pollution exposure observed at the national level, paralleling France’s situation where urban low-income groups disproportionately suffer from pollution (Le Thi, Suarez Castillo and Costemalle, 2023; Champalaune, 2020; Salesse et al., 2022). The spatial segregation of high and low-income populations in French urban and suburban areas further intensifies this phenomenon (Aerts, Chirazi and Cros, 2015).

Children’s sensitivity to air pollution exposure may also be influenced by inherent vulnerability factors (Deguen and Zmirou-Navier, 2010). Due to social inequalities in health, susceptibilities to air pollution may differ across standards of living, as children from lower-income families typically have poorer health at birth. Figure 4 highlights this disparity, showing that children from lowincome families are 1.5 times more likely to be born prematurely or with birth weights below 2.5 kg, representing a significant and noteworthy difference. These marked inequalities are also observed among children born at term, as detailed in Table A5 in the Appendix. Families with higher incomes tend to have more “healthy births”, defined as those with non-pathological admissions, and absence of principal diagnosis involving significant problems from the hospital discharge data.^4^ Of significant concern is the additional health care provided to children from lower-income families during their initial hospital stay at birth, especially for severe risks. Children from modest backgrounds born prematurely face a 1.77 times higher risk of low birth weight, a 1.48 times greater likelihood of needing neonatal intensive care, and a 1.75 times increased risk of undergoing a respiratory system X-ray.

**Figure 4:**
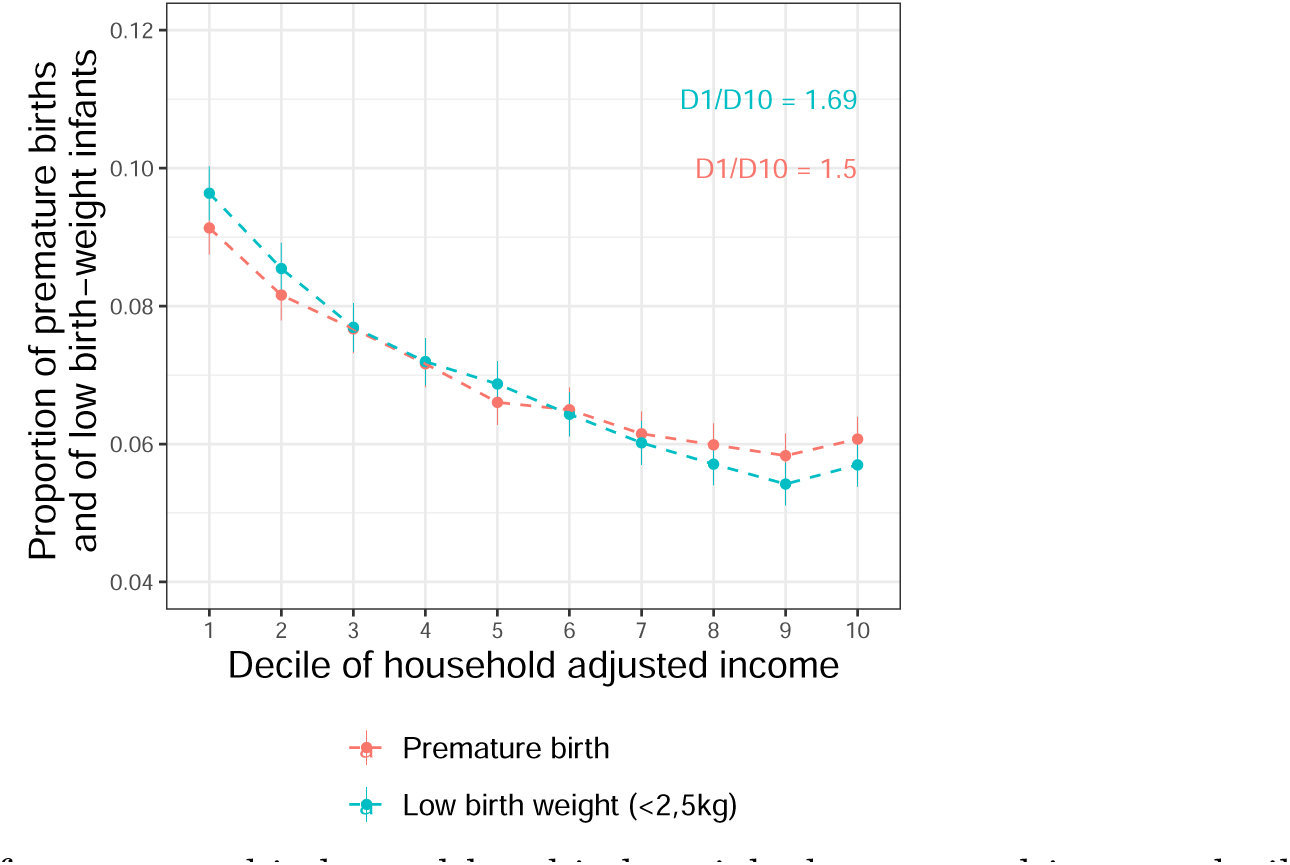
Share of premature births and low birth weight by parental income decile

Figure 5a demonstrates a significant correlation between parental income and the incidence of hospital stays for asthma in early childhood. Specifically, 1.9% of infants from the lowest income bracket require emergency hospital care, compared to only 1.2% from the highest income decile. This stark contrast underscores the pronounced health disparities across income groups in France, starting from birth. Perhaps more surprisingly, Figure 5a reveals that the administration of antiasthmatic medication increases with parental income up to the sixth decile. Given that asthma can be controlled with appropriate care, the higher levels of urgent asthma crises and lower levels of medication dispensed suggest insufficient treatments among lower-income groups. This is supported by the opposite patterns of increasing medication use and decreasing emergency admissions for related diseases up to the same income decile, implying that remedial investments are probably missing at the bottom of the parental income distribution. Medication usage across income levels mirrors the pattern observed in general practitioner and pediatrician visits, as depicted in Figure 5b. Public health guidelines mandate a specific number of doctor visits for all children, irrespective of their health status. From 2006 to 2018, nine such visits were required within a child’s first year, with full reimbursement from national health insurance, barring any extra fees charged by certain professionals.^5^ Doctor visits are more frequent among higher-income groups (up to the 7th decile). This may indicate differences in health care utilization rather than health status. Notably, lower-income families tend to consult general practitioners, whereas higher-income families more often visit pediatricians. These observations suggest disparities in both the quantity and quality of health care accessed by various income groups.

**Figure 5:**
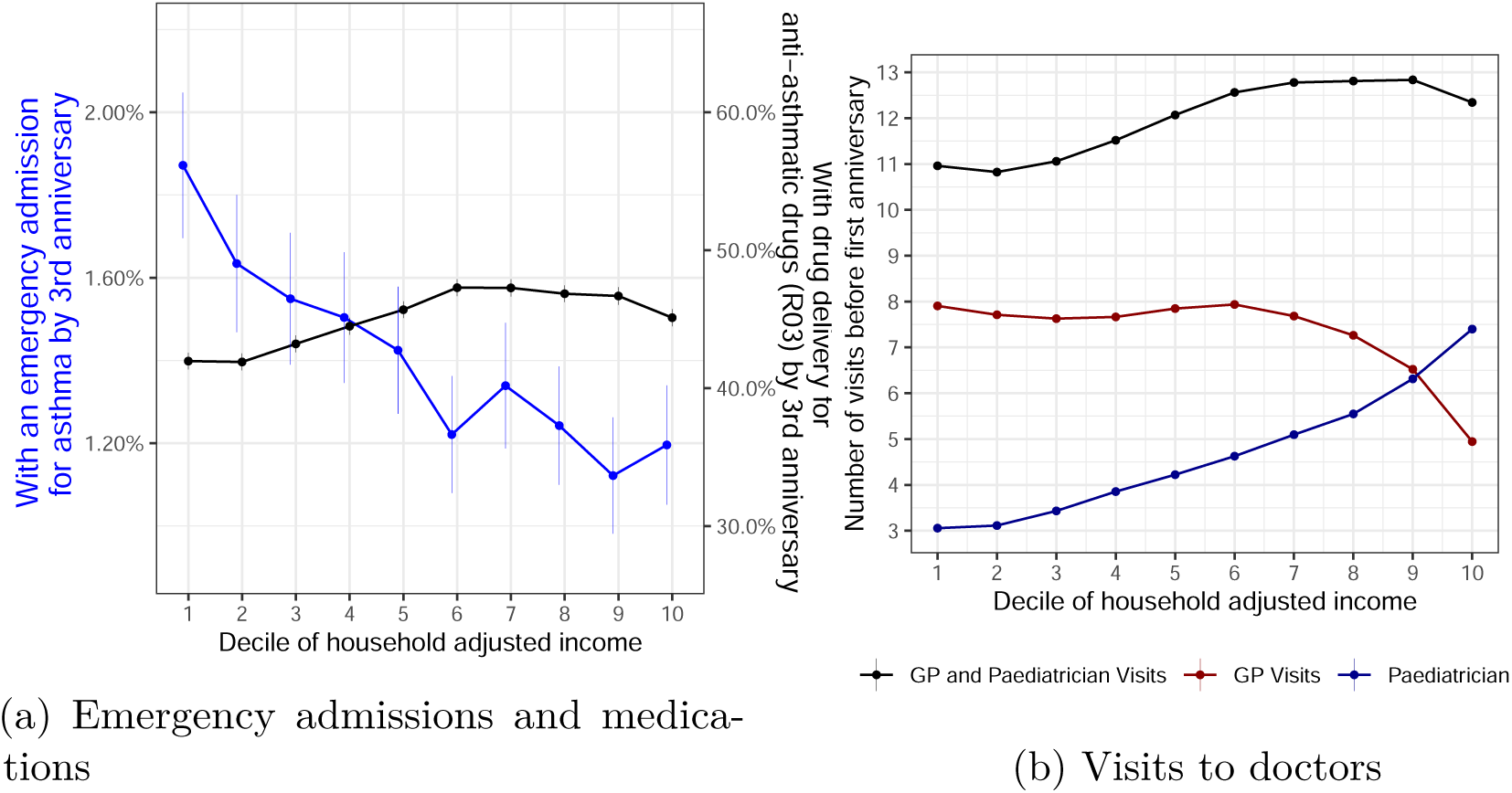
Emergency admissions for asthma, anti-asthma medications and visits to doctors (GP and pediatrician) by decile of parental income

## 3 Data

In this study, we combine several datasets at the municipality level: (i) air pollution data sourced from two distinct databases; (ii) atmospheric condition data extracted from a European reanalysis model; (iii) an extensive administrative dataset of a representative sample of the French population, with comprehensive individual data such as health care utilization and fiscal information.

Municipalities are delineated according to the 2017 administrative geography for *communes*, with *arrondissements* being used for major cities (Paris, Lyons, and Marseilles). These administrative units offer an adequate level of precision for the purpose of our study. Over half of these units have less than 500 inhabitants and occupy an area of less than 11 square kilometers (Chéron and Escapa, 2015), enabling a detailed study of exposure to air pollution.

### 3.1 Local Air Pollution

We use two reference data sources on local concentrations of PM2.5 in France at the annual level, to verify that our results are not sensitive to known discrepancies in large-scale estimates (Fowlie, Rubin and Walker, 2019). First, we use historical annual average data available for 2009-2020 at the municipality level in France. This database is produced by the French National Institute for Industrial Environment and Risks (Ineris). This dataset is the outcome of a statistical reconciliation process that combines measurements from monitoring stations with simulated data derived from an air quality model that uses emission inventories (Real et al., 2022). The second source is the Atmospheric Composition Analysis Group (ACAG) provides annual estimates of PM2.5 concentrations displayed on a one-kilometer grid that spans the European territory. These estimates are grounded on satellite observations and cover the years 2001-2018 (Hammer et al., 2020).

We evaluate the annual mean exposure to PM2.5 for children by merging these concentration data with location information, as detailed in Section 3.3. Although these datasets are available continuously over several years, one limitation is that they are only available for the calendar year. Therefore, as exposure in the first year of a child’s life starts on the date of birth, and not necessarily on the first of January, it is not possible to measure PM2.5 exposure for all children in our sample.

Figure 1 shows significant discrepancies in the average exposure of infants to PM2.5 between the two main data sources. The annual average exposure of infants in France sharply declined from 14-17 *µ*g/m^3^ in 2009 to 9-11 *µ*g/m^3^ by 2017. This decrease is explained by progress in all sectors, such as enhanced dust removal methods in industrial processes, improved efficiency of biomass combustion plants,^6^ and a reduction in primary particles emitted by diesel vehicle exhaust in large cities.^7^ In this context, an exposure variation of 0.10-0.15 *µ*g/m^3^ equates to 1% of the average annual exposure. This variation could represent a 1% sustained increase of the daily average concentration throughout a year, or alternatively, an amplification of the daily average concentration by 36.5% over a span of 10 days within a year. The quasi-experimental variations in annual average exposure examined in our study stem from a substantial short-term daily exposure surge, of about 30%, over a dozen of days with thermal inversions.

### 3.2 Local Weather Conditions

The UERRA datasets feature reanalysis data of atmospheric and surface climate variables, derived from the assimilation of historical data into a numerical weather model. The UERRA-HARMONIE reanalysis system offers hourly estimates of atmospheric variables at an 11km^2^ horizontal resolution, with 65 height levels, covering the period from 1961 to 2019 over Europe. This dataset, accessible on the Copernicus Climate Data Store, represents the highest resolution reanalysis dataset for Europe available at the time of our study, to the best of our knowledge.^8^ We derive municipality-level weather data by aligning each municipality’s centroid with its closest point on the UERRA grid.^9^ The weather-related control variables are established in accordance with the method described by Dechezleprêtre, Rivers and Stadler (2019). For each child, we acquire the following information for their first year of life: (1) the counts of the number of days during which the average daily temperature falls into one of 20 temperature bins, spanning the entire range of observed temperatures; (2) the counts of the number of days where the daily average wind speed is categorized into one of 12 wind speed bins, as delineated by the Beaufort wind scale; (3) the mean relative humidity experienced during their first year; and (4) the mean pressure throughout that year.

We classify a day with a thermal inversion when there is a positive difference between the daily mean temperatures at 500 meters and 15 meters above the surface level.^10^ We assign weather conditions, including thermal inversions, to each infant’s first year of life, considering their respective birth dates. Appendix A offers supplementary information pertaining to our weather data.

### 3.3 Health care use and socio-demographics

The *Echantillon Démographique Permanent* (EDP) is a longitudinal representative sample that tracks 4% of the French population across various administrative sources. Recently, it has been supplemented with National Health Insurance affiliation data, thereby creating the “EDP-Santé” data sample.

This rich dataset combines multiple data sources, including the census and several administrative sources for all individuals born on one of the first four days of each quarter, known as the “EDP individuals”, linked via a common identifier.^11^ Additionally, data from National Health Insurance (*Caisse Nationale d’Assurance Maladie*, CNAM) since 2008 provides detailed information on hospital admissions, drug prescriptions, and other health data.^12^ Our health data combines information about drug deliveries, inpatient birth stays, emergency admissions, and doctor visits. Income data from 2010 to 2016 is sourced from the fiscal database *Fidéli/Filosofi*, covering all EDP individuals and their household members, including parents, at the same address.

Our study encompasses 336,169 children born from 2008 to 2017. We track medication, specifically anti-asthma drugs (26% usage in the first year), delivered in city care and reimbursed by National Health Insurance over 2008-2018. These medications, classified under ATC code R03 for obstructive airway diseases,^13^ are observed alongside doctor consultations or visits resulting in individualized reimbursements. Data excludes consultations from the Maternal and Child Protection (*Protection maternelle et infantile*, PMI), with 15% of children under six having at least one consultation in 2012 (Amar and Borderies, 2015).

We use hospital discharge data from the PMSI (*Programme de Médicalisation des Systèmes d’Information*) that includes all hospital stays (inpatient hospital admissions) linked to EDP individuals from 2008 to 2018. Emergency admissions for asthma and bronchiolitis in infants’ first three years are identified using ICD-10 codes J45-J46 and J21 for the principal diagnosis encoded by the physician.^14^ These admissions exclude non-hospitalized emergency room visits, representing severe cases. The diagnosis used is determined at the end of the patient’s stay and justifies the hospital admission. Emergency admissions for bronchiolitis are considered before age two, and for asthma before age three, following Santé Publique France and Haute Autorité de Santé guidelines.^15^ and the Haute Autorité de Santé.^16^ The health data is further detailed in Appendix A.

We identify birth hospital stays for 85% of EDP infants using the methodology based on perinatal statistics.^17^ For a more comprehensive analysis, we use subset of (DREES) for measuring health inequalities (Dubost and Leduc, 2020) 235,000 EDP children, for whom we have both household income data from *FidéliFilosofi* and detailed birth health information. Parental income is calculated as adjusted disposable income, combining earnings, self-employment income, capital income, and social transfers, minus direct taxes, adjusted for household size (Blanpain, 2019).^18^ We rank infants by household income percentile within their birth cohort for the first three years, and assign them to income groups using the average income percentile across these years.

## 4 Empirical Strategy

Our objective is to estimate the causal effect of an increase in exposure to air pollution on anti-asthmatic drugs deliveries and emergency admissions during the first years of life. Identifying the causal effects of pollution exposure on children’s health care use is however challenging due to several factors. First, pollution is not randomly distributed across space, as it is often associated with the proximity to dense urban areas. These areas have specific populations with varying baseline health, health behaviors, and access to health care.^19^ Second, even within a specific location, pollution is not randomly assigned over time. It tends to be correlated with local economic conditions, which can influence or be associated with infant health outcomes.^20^ Third, individual measures of pollution exposure are subject to errors.

This section presents our identification strategy, the econometric models used to estimate the average treatment effects, and our approach to investigate heterogeneous treatment effects.

### 4.1 Linking Air Pollution to Health Care Use

We tackle endogeneity by isolating and designing a quasi-experimental air pollution binary shock induced by variations in local thermal inversion exposure among children, factoring in their birth location and time. This approach allows us to create distinct “more exposed” (treated) and “less exposed” (control) cohorts for comparison.

We opt for a reduced-form IV, often referred to as ITT, for two primary reasons. The first is methodological: the approach developed by Chernozhukov et al. (2023) used in the heterogeneity analysis requires a binary treatment variable and does not accommodate instrumental variables. We hence closely follow the approach in Deryugina et al. (2019) by designing a binary treatment from a plausibly exogenous variable. The second reason pertains to data constraints: the annual aggregation of pollution data limits the first stage of our analysis to only children born in January, resulting in the exclusion of approximately 75% of the sample. Consequently, while we also present results using a two-stage least squares (2SLS) method with this restricted first stage in the appendix, our primary focus is on the ITT approach.^21^

#### Identification strategy

Our identification strategy is to compare children who have been exposed to an above-normal number of thermal inversions during their first year to others, where the “normal level” varies across locations and years. The key identifying assumption is that after controlling for municipality fixed-effects, year fixed-effects, and weather variables, the *local changes* in the number of thermal inversions are unrelated to changes in the health outcomes of children except through their influence on air pollution. In this context, we interpret our binary treatment as a positive shock of air pollution exposure. We use this treatment for both estimating average treatment effects and conducting the heterogeneity analysis.

The binary treatment variable *T_i_* serves as a quasi-random assignment to higher levels of air pollution exposure. It assigns children either to the more exposed or less exposed groups based on their exposure to temporary surges in air pollution, as measured by an above-average number of days with thermal inversions before their first anniversary. The reference level of number of thermal inversion days for each infant *i*, denoted *N̅_i_*, is calculated at the municipality *c* and year *t* level. This baseline accounts for regional and temporal variations, including geographical factors like topography and annual fluctuations potentially linked to climate change. Figure 6 illustrates the relationship between thermal inversions and topographical characteristics, which are irrelevant to our identification strategy. Appendix B.1 is devoted to our model of *N̅_i_*.

**Figure 6:**
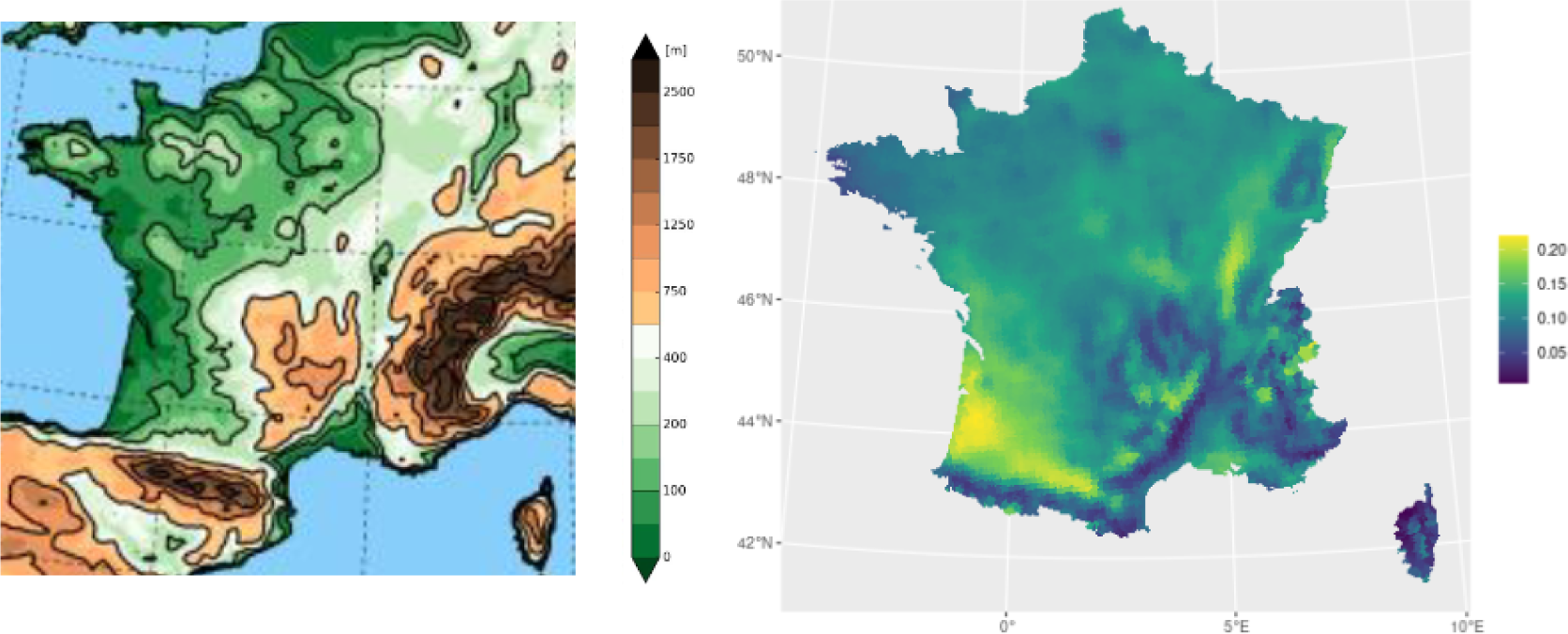
Topography in the UERRA model (source: UERRA user guide) and annual share of days with thermal inversions UERRA 1999-2017 (right).

For each infant *i*, we calculate the exposure to *n_i_* = *N_i_* − *N̅_i_* additional days of thermal inversion compared to the local long-term average *N̅_i_*, which varies across birth year *t* and municipality *c*. Here, *N_i_* represents the actual number of days with thermal inversion experienced by child *i* in their first year. We define a cohort as exposed when the number of days with local thermal inversion during the child’s first year of life exceeds the reference level by a threshold value denoted as *n >* 0. The binary treatment variable is formally defined as follows:

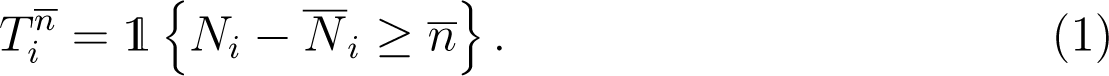

We also conduct our analysis using *T_i_* as an instrumental variable for PM2.5, which yield estimates of average treatment effects closely aligned with our reduced-form results.^22^ However, we have made a deliberate choice not to emphasize these results in the main specifications of the paper and instead focus on the ITT for the reasons mentioned above.

#### Econometric specifications

We estimate the average treatment effects on health outcomes with the following model

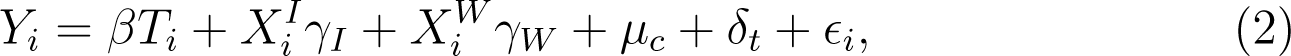

where the dependent variable *Y_i_* is the health care outcome of interest for child *i*, born in municipality *c*. We control for individual-level variables *X^I^_i_*, weather variables *X^W^_i_* , as well as municipality fixed effects *µ_c_* to control for local and time-invariant determinants of health and pollution, and birth-year fixed effects *δ_t_* to control for unobserved time-varying shocks common to children born in the same year *t*. *T_i_* is constructed to represent that child *i* is exposed to an air pollution shock before their first anniversary, which is mediated by exposure to an *above normal* number of thermal inversions. This approach ensures that the air pollution exposure is arguably exogenous to the child’s unobservable characteristics, as will be demonstrated shortly.

At the individual level, we control for a very large set of variables aimed at capturing observed heterogeneity: gender, parental income (introduced in decile and linearly), mother’s characteristics (age and an indicator for being born abroad), gestational age (as an indicator for premature birth and linearly), birth weight (low birth indicator and linearly), as well as six other health indicators derived from the hospital stay at birth.^23^ In addition, *X^W^_i_* includes an extensive range of weather conditions characterizing the child’s exposure during the first year, i.e. the number of days in each of 20 temperature bins, the number of days in each of 12 wind strength bins, and second-order polynomials for average pressure and humidity.

The parameter of interest is *β*, interpreted as the average treatment effect associated with the quasi-experimental binary shock of exposure to air pollution *T_i_* ∈ {0, 1}. All standard errors are clustered at the UERRA grid level, the geographical level of measure of the treatment status.

#### Quasi-experimental pollution exposure shocks

The estimates derived from (2) can be seen as the ITT effects, capturing the reduced-form impact of the treatment variable *T_i_* which serves as an instrumental variable used to quasi-randomly assign infants to air pollution shocks. We investigate the threshold choice *n* using the corresponding first-stage regression

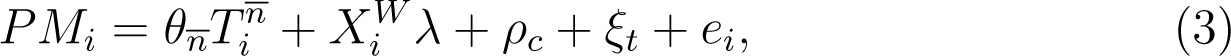

where *PM_i_* measures concentrations of fine particulate matter for child *i* given its municipality *c* of residence and year of birth *t*, varying *n̄*. We control for weather variables, *X^W^* , as well as municipality and year fixed effects, respectively *ρ_c_* and *ξ_t_*. Recall that PM2.5 data is only available at the annual level. Therefore, we estimate (3) for children born in January, whose first year coincides with the calendar year. This restriction reduces the sample size by four. Additionally, including controls *X^I^* reduces the sample by approximately 40% due to missing values.

In our analysis, we opt for the threshold *n̄* = 7, which corresponds to an intermediate pollution shock equivalent to approximately one standard deviation of *n_i_* (sd = 7.7). This approach leads to categorizing 14% of the sample as the exposed group. Figure 7 displays the estimation results for *θ*, showing a dose-response relationship between PM2.5 levels and local changes in thermal inversions from both reference data sources on pollution (ACAG and Ineris).^24^ We further evaluate how this quasi-experimental shock impacts the annual number of thermal inversions experienced by children with the following specification

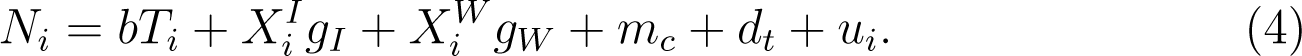

**Figure 7:**
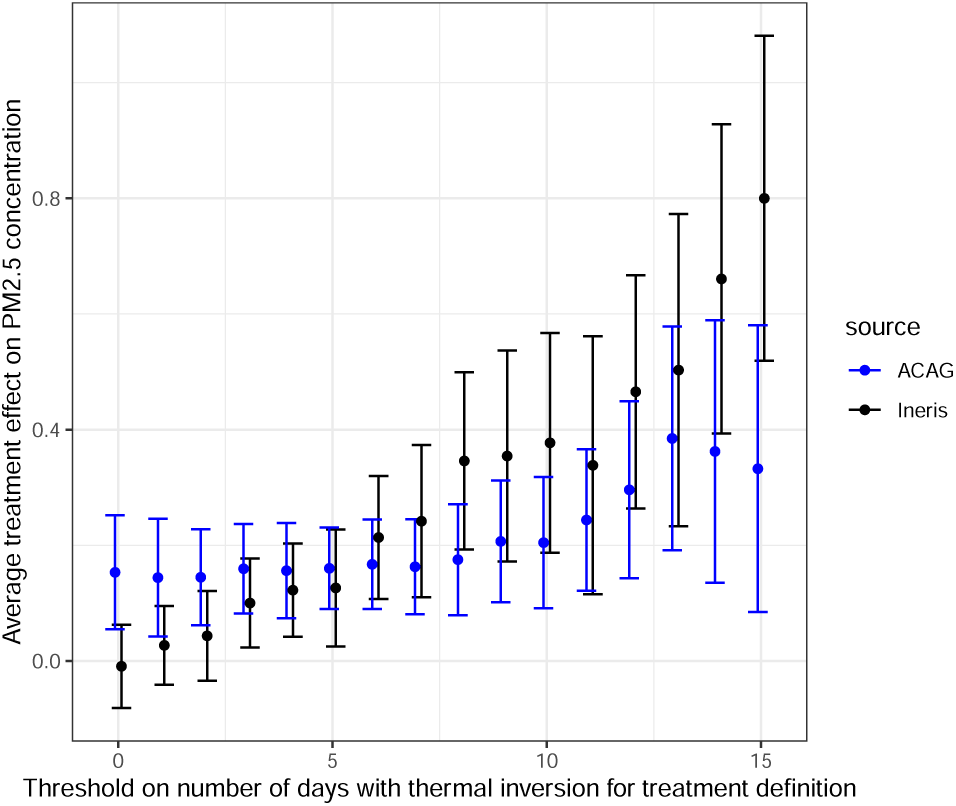
Days with thermal inversion from birth to first anniversary and PM2.5 Exposure in first year. *Notes: Infants born in January from the primary sample. θ_n_ estimates from* (3) *for increasing values of threshold n*.

Table 1 reports the corresponding estimates from (3) and (4). According to column (1), children in the exposed group, on average, experience an additional 10.8 days of thermal inversion before their first anniversary. Depending on the PM2.5 source, this corresponds to an increase in annual average exposure by either 0.12 or 0.24 *µ*g/m^3^. The estimates in Table 1 remain robust when including *X^I^* in column (2) despite the reduced sample size. However, for the ACAG source, the coefficient is slightly reduced and not significant at the 5% level. Columns (3) and (4) report estimates when excluding children who are intermediately treated, i.e. ∀*n_i_* ∈ [0; *n*]. In this case, the treated and control groups differ by almost 15 days of thermal inversions, leading to a larger PM2.5 exposure difference of approximately 0.3 *µ*g/m^3^ in both sources.^25^

**Table 1:** Design of the quasi-experimental treatment and exposure to air pollution: when days with thermal inversion in first year exceed by *n* = 7 days the prediction

| *Outcome* | All Children |  | Excluding those<br>intermediately treated |  |
| --- | --- | --- | --- | --- |
|  | (1) | (2) | (3) | (4) |
| *Additional days with thermal inversion in first year* |  |  |  |  |
|  | 10.79 | 10.60 | 14.71 | 14.65 |
|  | [10.48;11.11] | [10.32;10.89] | [14.31;15.11] | [14.33;14.97] |
| $X^I$ | no | yes | no | yes |
| Sample size | 336169 | 217859 | 235380 | 147042 |
| *Additional exposure to PM2.5 (ACAG source) in first year for those born in January* |  |  |  |  |
|  | 0.12 | 0.055 | 0.28 | 0.21 |
|  | [0.022;0.22] | [-0.044;0.15] | [0.17;0.40] | [0.075;0.35] |
| $X^I$ | no | yes | no | yes |
| Sample size | 82164 | 49703 | 49133 | 29732 |
| *Additional exposure to PM2.5 (Ineris source) in first year for those born in January* |  |  |  |  |
|  | 0.24 | 0.23 | 0.29 | 0.23 |
|  | [0.12;0.37] | [0.085;0.37] | [0.14;0.45] | [0.043;0.43] |
| $X^I$ | no | yes | no | yes |
| Sample size | 73354 | 49214 | 44530 | 29462 |

Notes: This Table presents the results of twelve distinct regressions, with three outcomes in rows and four specifications in columns. Each outcome is regressed on the binary exposure dummy $T_i = 1\{\hat{n} > \bar{n} = 7\}$ , on ground-level weather controls, with or without individual characteristics $X^I$ , and with municipality and year fixed effects. Standard errors are clustered at the UERRA grid level.

We conduct a comparison exercise to validate the assumption that the exposure dummy provides a reliable exogenous variation of air pollution. In (2), we replace *Y_i_* with infant characteristics *X^I^*, excluding them from the set of explanatory variables. The resulting coefficients in Table 2 represents the conditional correlation between the treatment status and child characteristics. These results indicate a high level of comparability between treated and non-treated births regarding children’s characteristics, both unconditionally (columns 2 and 3) and conditionally (columns 4 and 5). This comparability provides credibility to using the exposure dummy based on thermal inversions as a reliable proxy for as-good-as-random variations in air pollution.^26^ Additionally, we demonstrate that the temporal variation in children’s exposure to thermal inversion days across urban areas is free from specific trends in Figure B4 in Appendix B. This ensures that our analysis is not influenced by particular temporal biases.

**Table 2:**
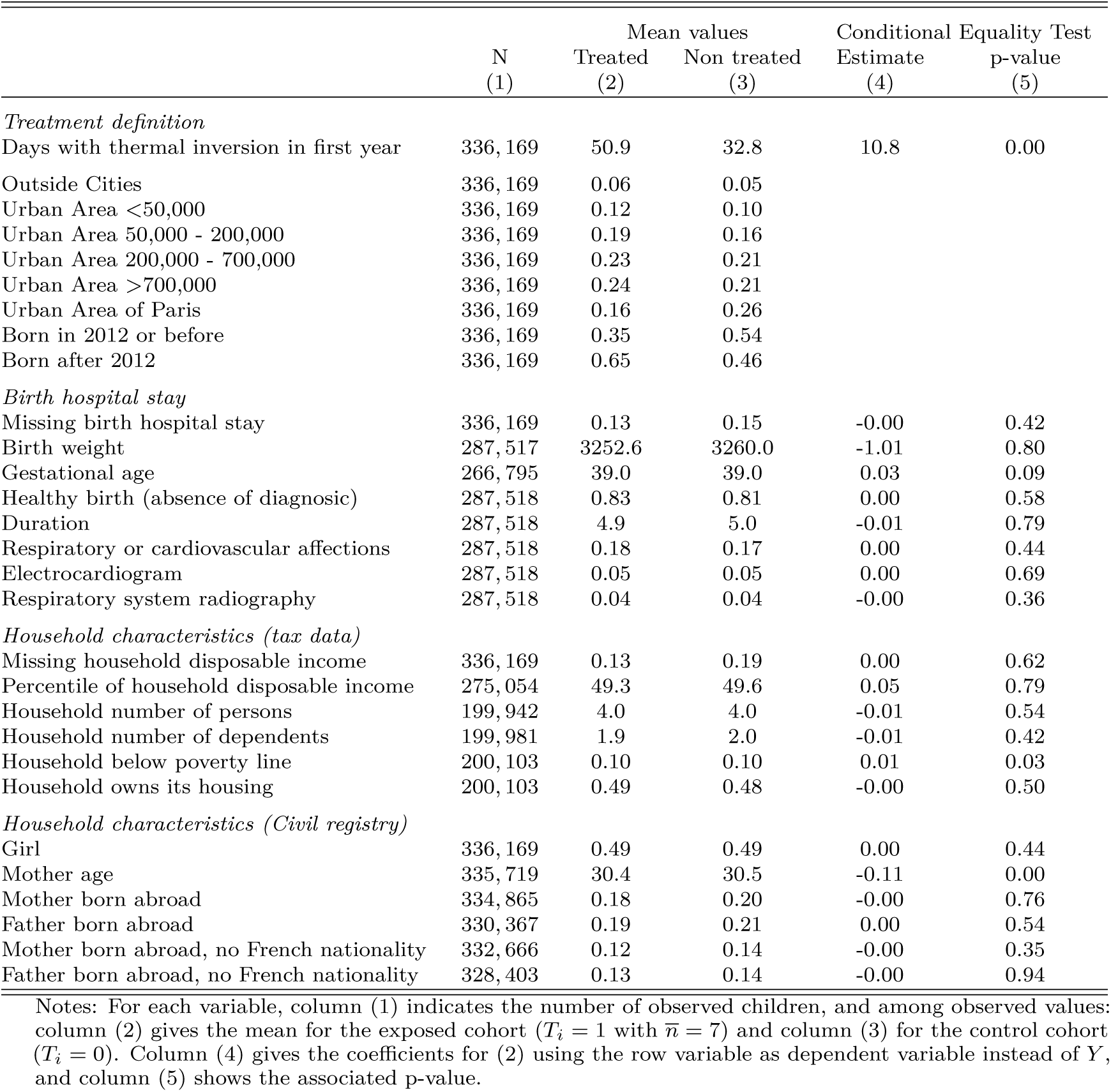
Characteristics of the sample and birth comparability across treated and non treated infants.

### 4.2 Uncovering Inequalities with Machine Learning

The primary purpose of our study is to document children’s health inequalities related to air pollution in three ways. We aim at: (a) confirming that children react heterogeneously to air pollution; (b) quantifying the distribution of negative effects across children; and (c) drawing a portrait of the most affected children, in terms of parental income and vulnerability factors.

Our descriptive evidence on exposure and health care use suggests that the effects of air pollution may be vastly heterogeneous across children of different income groups. However, adding interactions terms to model (2) would provide a limited portrait of the most vulnerable children by choosing ex-ante a limited number of groups to compare based on some variables. Therefore, we study the heterogeneous treatment effects of our binary quasi-experimental shock using the generic machine learning approach developed in Chernozhukov et al. (2023). We propose an intuitive description of this procedure below and provide all the technical details in Appendix C.

We maintain the same exposure groups (treated and control) as described above, but we introduce propensity scores to replace the large set of municipality fixed effects for computational reasons, and also to address potentially remaining group imbalances.^27^ We search for heterogeneity using all explanatory variables listed in Table C1, denoted *Z_i_*, i.e. the same variables *X_i_^I^* used in (2) in addition to extra local characteristics: a measure of local concentrations of PM2.5 before birth, a measure of accessibility to GPs and to pediatricians, and the type of urban areas as depicted in Figure A3. To predict the health care use of children, we train and tune two machine learning algorithms using 100 random splits of the data, where each split randomly assigns half of the data to a main sample and the other half to an auxiliary sample. The algorithms aim to predict the health outcome of infant *i* conditional on all observable covariates *Z_i_*, separately for the infants in the exposed group and the control group, and use only the auxiliary samples for both training and tuning. We use these machine learning predictions, denoted *Ŷ^T^* (*Z_i_*) and *Ŷ^C^* (*Z_i_*), respectively, to form proxy predictors in the main sample of: (1) the outcome of interest of each infant in absence of pollution shock *b*_0_(*Z_i_*) = E[*Y_i_*|*Z_i_, T_i_* = 0] with *b̂*(*Z_i_*) = *Ŷ^C^* (*Z_i_*), and (2) the conditional average treatment effect (CATE) of each infant *s*_0_(*Z_i_*) = E[*Y_i_*|*Z_i_, T_i_* = 1] − E[*Y_i_*|*Z_i_, T_i_* = 0] using *Ŝ*(*Z_i_*) = *Ŷ^T^* (*Z_i_*) − *Ŷ^C^* (*Z_i_*).

The output *Ŝ*(*Z_i_*) is used as a single index capturing each infant’s sensitivity to the air pollution quasi-experimental variation. This index represents the likelihood of infant *i*, with characteristics *Z_i_*, to need healthcare services for respiratory issues resulting from increased air pollution exposure in their first year. It is possible to conduct valid inference about important features of the true CATE *s*_0_(*Z_i_*), even when *Ŝ*(*Z_i_*) is a biased estimate, by using the main samples across all data splits (Chernozhukov et al., 2023). The two proxy predictors and the estimated propensity scores are used to address our three aims by: (a) estimating the Best Linear Predictor of the conditional average treatment effect *s*_0_(*Z_i_*) (BLP); (b) estimating the Group-Average Treatment Effects by sensibility groups (GATES); and (c) performing a classification analysis to describe the vulnerability groups (CLAN, for classification analysis).

The BLP of *s*_0_(*z*) is an affine function *β̂*_1_ + *β̂*_2_*Ŝ*(*z*) where *Ŝ*(*z*) acts as a proxy for treatment effect heterogeneity. We estimate it using Ordinary Least Squares (OLS) on the main sample, projecting the outcome *Y_i_* onto the treatment interacted with the proxy variable *Ŝ*(*Z_i_*) and incorporating additional control variables. Typically, this interaction method is used to explore heterogeneity across specific dimensions of *Z_i_*. However, in our approach, the heterogeneity dimension is not predetermined by the researcher but is instead determined through *Ŝ*(.), by dedicating half of the sample to this learning process. The estimated coefficient *β̂*_2_ is a consistent estimate of the correlation between the treatment sensitivity, as measured by *Ŝ*(*z*), and the true CATE.^28^ Testing the null hypothesis *β*_2_ = 0 hence provides a way to investigate the presence of heterogeneous treatment effects, i.e. that *s*_0_ varies with *Z*. Rejecting *β*_2_ = 0 means both that *s*_0_ varies with *Z* and that *Ŝ*(·) is a relevant proxy predictor of the heterogeneity. If not rejected, it could be that *s*_0_ does not vary with *Z* or that *Ŝ*(·) does not capture well the true heterogeneity.

We create four groups *k* = 1, 2, 3, 4 based on increasing predicted average treatment effects, using the 50th, 75th, and 90th quantiles of *Ŝ*(*Z_i_*). We estimate the GATES by OLS, under the same rationale: using half of the sample to form sensitivity groups facilitates estimating interactions with the treatment dummy in the main sample to represent average effect for each group. The resulting estimates *γ̂_k_*’s corresponds to the expected *s*_0_(*z*) for each group *k*. By construction, the GATES are expected to satisfy *γ̂*_1_ ≤ *γ̂*_2_ ≤ *γ̂*_3_ ≤ *γ̂*_4_ reflecting the average treatment effects correlating to each group’s sensitivity. This approach provides valuable insights into the distribution of the negative health impacts of the quasi-experimental pollution shock across children in our data. Absence of heterogeneity or an ineffective proxy would result in statistically indistinguishable GATES across groups.

Finally, we investigate the compositions of these groups defined in the GATES using the vector of CLAN parameters containing the average characteristics of each group *k*, denoted *δ_k_*, along the multiple dimensions of *Z*. This approach hence allows drawing a portrait of the different groups, in particular the most affected.

The parameters of interest are therefore *β*_1_, *β*_2_, *γ_k_*’s, and *δ_k_*’s, which are separately estimated for each main sample across all 100 data splits. We report their median across the splits, and their confidence intervals of coverage 1 − *α*, as calculated by Chernozhukov et al. (2023) as the median across the splits of CIs with coverage 1 − *α/*2, which is rather conservative.^29^

## 5 Results

The next two subsections include the main results: average effects and heterogeneity analysis. The appendices provide further insights, including an examination of emergency admission avoidability (A.1.2), a placebo test (B1), IV estimates (B.2), additional outcomes (B.4), robustness regarding seasonality and quarter-of-birth (B.1 and B.3), and an additional heterogeneity analysis including measures of healthcare accessibility (C1).

### 5.1 Average effects on health care use

We first estimate the average effects of the air pollution shock on antiasthmatic drug deliveries and emergency admissions. Table 3 reports the parameter estimates and standard errors associated with *T^n̂=7^_i_* from (2). The first two columns report the average treatment effect for a baseline exposure shock, where the exposed group consists of infants with *n_i_ > n̂* = 7, and the control group comprises infants with *n_i_* ≤ *n̂* = 7. The last two columns show the same specifications but omit infants with intermediate exposure, comparing infants with *n_i_ > n* to infants with *n_i_ <* 0. In both instances, the second column limits the sample to infants with (non-missing) individual characteristics *X^I^* for robustness. The results are largely insensitive to the inclusion of *X^I^*, aligning with the absence of correlation between an infant’s exposure status and its characteristics, as shown in Table 2.

**Table 3:** Average treatment effect of the quasi-experimental air pollution shock

| *Outcome* | All Children |  | Excluding those intermediately treated |  |
| --- | --- | --- | --- | --- |
|  | (1) | (2) | (3) | (4) |
| *Having drug delivery for obstructive airway diseases before first anniversary* |  |  |  |  |
|  | 0.003 | 0.003 | 0.008 | 0.006 |
|  | [-0.003;0.009] | [-0.004;0.010] | [0.001;0.014] | [-0.002;0.014] |
| $X^I$ | no | yes | no | yes |
| Sample size | 336169 | 217859 | 235380 | 147042 |
| *Hospital emergency admissions: bronchiolitis or asthma before third anniversary* |  |  |  |  |
|  | 0.005 | 0.007 | 0.004 | 0.006 |
|  | [0.002;0.009] | [0.003;0.011] | [0.000;0.008] | [0.002;0.011] |
| $X^I$ | no | yes | no | yes |
| Sample size | 246075 | 189722 | 163071 | 124262 |
| *Hospital emergency admissions: asthma before third anniversary* |  |  |  |  |
|  | 0.002 | 0.003 | 0.002 | 0.003 |
|  | [0.000;0.005] | [0.001;0.006] | [-0.000;0.005] | [-0.000;0.005] |
| $X^I$ | no | yes | no | yes |
| Sample size | 246075 | 189722 | 163071 | 124262 |
| *Hospital emergency admissions: bronchiolitis before second anniversary* |  |  |  |  |
|  | 0.003 | 0.003 | 0.004 | 0.004 |
|  | [0.000;0.006] | [-0.000;0.006] | [0.001;0.007] | [0.000;0.008] |
| $X^I$ | no | yes | no | yes |
| Sample size | 277271 | 207825 | 188460 | 139968 |

*Notes:* This table presents the results of 16 distinct regressions, with 4 outcomes in rows and 4 specifications in columns. Each outcome is regressed on the binary treatment designed to capture a positive variation in air pollution exposure, $T_i = 1\{\hat{n} > \bar{n} = 7\}$ , quantifying whether a child was over-exposed to thermal inversion in its first year, on ground-level weather controls, with or without individual characteristics $X^I$ , municipality and year fixed effects. Standard errors are clustered at the UERRA grid level.

Exposure to an air pollution shock, corresponding to an additional 0.1 to 0.2 *µg/m*^3^ in the first-year average exposure to PM2.5 (*n̂* > *n̂*), causally increases the risk of infants being admitted to the emergency room for asthma (resp. bronchiolitis) before the third anniversary by 0.2 percentage points (resp. 0.3). This corresponds to 14% (resp. 8%) of the baseline risk of 1.4% (resp. 3.6%). Notably, the risk of receiving anti-asthma medications before the first anniversary significantly increases only when treated and controls have larger exposure differences (column 3), corresponding to an additional 0.3 *µg/m*^3^ in first-year average PM2.5 exposure. Note that, given the disparity between PM2.5 data sources and the variety of pollutants affected by thermal inversion, these estimates provide evidence of a causal link rather than a precise quantification.^30^

Due to the somewhat arbitrary nature of selecting the threshold, we also present the estimates for various choices in Figure 8. We observe dose-response relationships like in Figure 7 for PM2.5 concentrations, suggesting that our treatment design effectively captures the consequences of air pollution on infants’ health care use. Specifically, the risk of receiving antiasthmatic drugs before the first anniversary is significantly increased at the 5% level starting at *n̂* = 10, while the risk of emergency admission for asthma or bronchiolitis and the number of visits to doctors are significantly increased at the 5% level starting at *n̂* = 5. The average treatment effect for medication becomes more pronounced when excluding moderately treated infants, whereas the point estimates for emergency admissions barely change. This suggests a stronger dose-response relationship for medication and possibly a threshold effect for emergency admissions.

**Figure 8:**
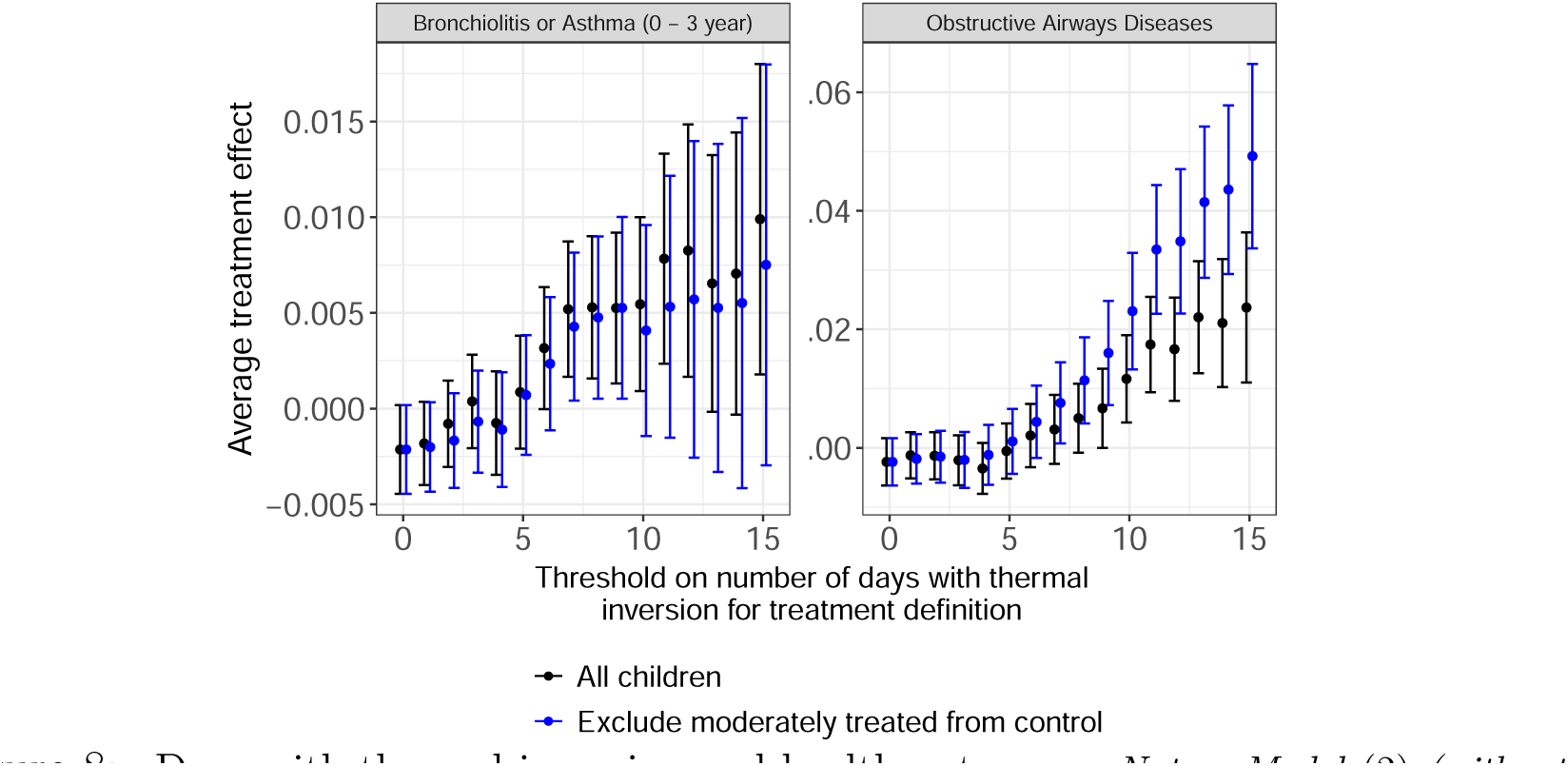
Days with thermal inversion and health outcomes. *Notes: Model* (2) *(without X^I^) for increasing values of n*.

In a placebo test, we identify a cohort of placebo-treated infants residing in municipalities that experience an increase in thermal inversions (*n̂* > *n̂*) after their healthcare consumption period. These children belong to areas where an unusual number of thermal inversions occur in the year following their n-th anniversary, hence the exposure occurs after the considered period, with the outcome measured from birth to their n-th anniversary. We estimate (2), without individual controls to maintain a large sample size, among never-treated children from birth to their n-th anniversary. The results, reported in Table B1 in Appendix B, show no significant effects from this placebo treatment, providing evidence for the credibility of our analysis.

### 5.2 Heterogeneity of the effects

#### Evidence of heterogeneity

Table 4 presents the best linear predictor of the CATE for each outcome. In almost all cases, we reject homogeneity at the 5% level, based on the p-values associated with the coefficient *β̂*_2_, which captures the correlation between the CATE and the proxy. The most relevant proxy, in terms of their correlation with the CATE, is found for bronchiolitis emergency admissions, with *β̂*_2_ estimated at 0.33 (p-value < 0.001). For antiasthmatic medication in the first year, *β̂*_2_ is estimated at 0.11 (p-value = 0.003). For asthma emergency admissions before the third anniversary, *β̂*_2_ is estimated at 0.162 (p-value = 0.001). These results indicate that our algorithms effectively capture significant heterogeneity in treatment effects.^31^

**Table 4:** Testing for heterogeneity and proxy relevance: Best Linear Predictor

|  |  |
| --- | --- |
| *Having drug delivery for obstructive airway diseases before first anniversary* |  |
| ATE($\hat{\beta}_1$) | 0.006 [-0.003;0.014] p=0.201 |
| HTE($\hat{\beta}_2$) | 0.114 [0.038;0.190] p=0.003 |
| *Hospital emergency admissions for bronchiolitis or asthma before third anniversary* |  |
| ATE($\hat{\beta}_1$) | 0.004 [-0.000;0.009] p=0.066 |
| HTE($\hat{\beta}_2$) | 0.136 [0.038;0.237] p=0.007 |
| *Hospital emergency admissions for asthma before third anniversary* |  |
| ATE($\hat{\beta}_1$) | 0.002 [-0.000;0.005] p=0.067 |
| HTE($\hat{\beta}_2$) | 0.162 [0.063;0.258] p=0.001 |
| *Hospital emergency admissions for bronchiolitis before second anniversary* |  |
| ATE($\hat{\beta}_1$) | 0.003 [-0.001;0.006] p=0.141 |
| HTE($\hat{\beta}_2$) | 0.325 [0.248;0.401] p=0.000 |

Notes: Medians over 100 splits. The reported confidence intervals are the median across the splits of CIs with coverage $1 - \alpha$ with $\alpha = 0.05$ . Associated p-values are computed as two times the median across the splits of the p-values that the parameter is equal to zero against the two-sided alternative, and could be divided by two under a mild assumption (see Appendix C).

We also experimented with another choice of *n*. In general, the machine learning-derived heterogeneity correlates better with the ground truth for an intermediate treatment level compared to high treatment levels because the latter substantially reduces the size of the treatment group. For instance, the intermediate threshold *n̂* = 7 yields a treatment group with about 14% of infants compared to about 8% for *n̂* = 10. The results are nevertheless qualitatively similar, though less precise.

#### Concentrated effects

For anti-asthma drug deliveries and emergency admissions for bronchiolitis, we find compelling evidence that the pollution exposure effect is concentrated in 10% of the infants, which consists of infants between the 90th and 100th percentile of the proxy predictor for each outcome, as shown in Table 5 and illustrated in Figure 9. Exposed infants in these groups have a 2.4 p.p. (p-value = 0.053) higher probability of using antiasthmatic medication in the first year and a 1.7 p.p. (p-value = 0.002) higher probability of emergency admission for bronchiolitis.

**Table 5:** Average effect by sensitivity group

|  |  |  |
| --- | --- | --- |
| *Having a drug delivery for obstructive airway diseases before first anniversary* |  |  |
| GATE($\hat{\gamma}_1$), bottom 50% | -0.001 [-0.012;0.010] | p=0.809 |
| GATE($\hat{\gamma}_2$), 50th-75th percentile | 0.007 [-0.007;0.022] | p=0.313 |
| GATE($\hat{\gamma}_3$), 75th-90th percentile | 0.009 [-0.010;0.028] | p=0.206 |
| GATE($\hat{\gamma}_4$), top 10% | 0.023 [-0.001;0.046] | p=0.053 |
| GATE($\hat{\gamma}_4$) - GATE($\hat{\gamma}_1$) | 0.024 [-0.001;0.049] | p=0.065 |
| *Hospital emergency admissions for bronchiolitis before second anniversary* |  |  |
| GATE($\hat{\gamma}_1$), bottom 50% | 0.000 [-0.004;0.005] | p=0.855 |
| GATE($\hat{\gamma}_2$), 50th-75th percentile | 0.002 [-0.005;0.008] | p=0.634 |
| GATE($\hat{\gamma}_3$), 75th-90th percentile | 0.001 [-0.008;0.009] | p=0.856 |
| GATE($\hat{\gamma}_4$), top 10% | 0.017 [0.006;0.028] | p=0.002 |
| GATE($\hat{\gamma}_4$) - GATE($\hat{\gamma}_1$) | 0.017 [0.006;0.029] | p=0.004 |

Notes: Medians over 100 splits. The reported confidence intervals are the median across the splits of CIs with coverage $1 - \alpha$ with $\alpha = 0.05$ . Associated p-values are computed as two times the median across the splits of the p-values that the parameter is equal to zero against the two-sided alternative, and could be divided by two under a mild assumption (see Appendix C).

**Figure 9:**
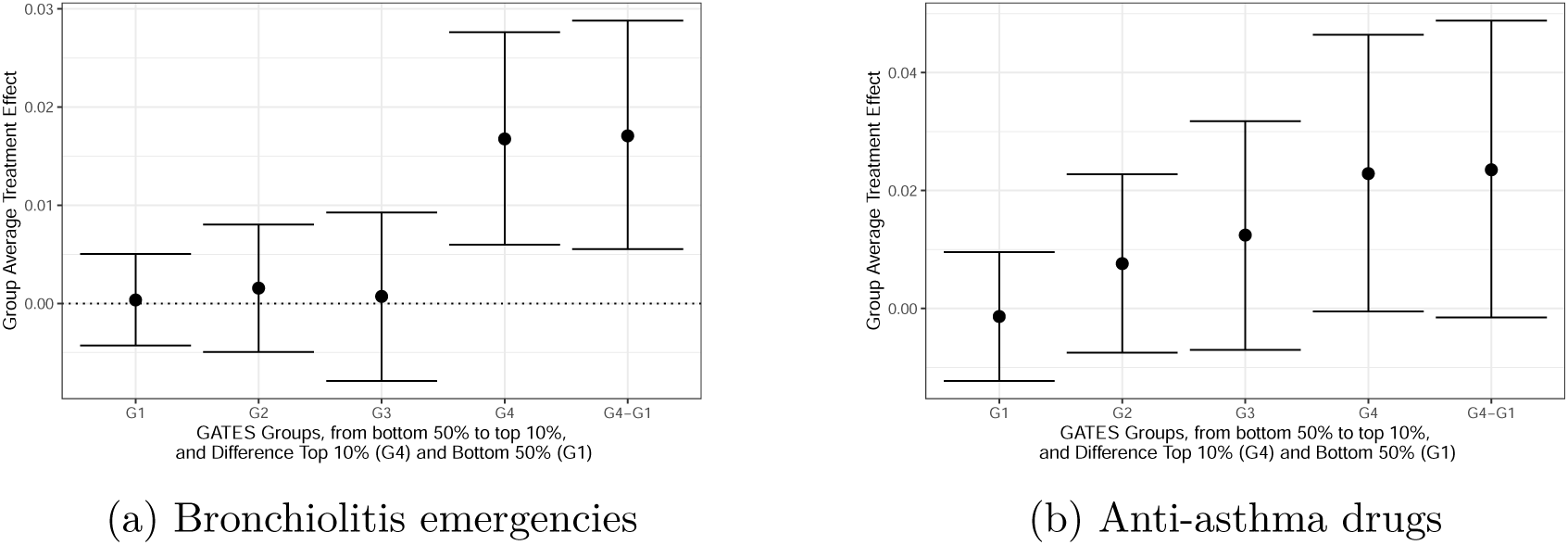
Average effect by sensitivity group. *Notes:* Medians over 100 splits. The reported confidence intervals are the median across the splits of CIs with coverage 1 − *α* with *α* = 0.05.Associated p-values are computed as two times the median across the splits of the p-values that the parameter is equal to zero against the two-sided alternative, and could be divided by two under a mild assumption (see Appendix C). The dashed lines report the ATE.

We observe progressive increases in the group average treatment effects for obstructive airways disease drugs, with significance observed primarily in the top decile. In comparison, the top 10%’s effects for bronchiolitis are more pronounced. In the third group (infants in the 75th to 90th percentile of predicted impacts), the parameter estimates are near zero and not significant. These results indicate that the significant effects of air pollution exposure are largely concentrated in the most affected 10% of infants, diminishing quickly for the less sensitive groups.

#### Portraying the most vulnerable children

Table 6 shows the CLANs for the two outcomes identified by the GATES, focusing on the top 10% of most affected infants. This table describes the mean characteristics of this group compared to the bottom 50%. The results for both outcomes reveal the following patterns. Primarily, infants with pre-existing health challenges (such as preterm birth, low birth weight, cardio-respiratory pathology, electrocardiogram abnormalities, and respiratory X-rays at birth) are more adversely affected by excessive exposure, as reflected in both outcomes. The top 10% most affected infants for emergency admissions for bronchiolitis are particularly vulnerable. For instance, they are almost three times more likely of being born prematurely (18.7% risk compared to 5.9% for the 50% least affected) and have an 18.9% risk of low birth weight. In contrast, the differences between the 10% most affected and 50% least affected in terms of medication are relatively modest, showing only a 2.6 p.p. difference. Regarding emergency admissions for bronchiolitis, the most affected infants are often economically disadvantaged on several dimensions. Specifically, the bottom 10% in parental income represents 17.4% of this group, and beneficiaries of the universal health coverage are disproportionately represented as discussed below. For anti-asthma medications, there is only a slight over-representation of the lowest income decile (11.1% in the most affected versus 9.4% in the least affected). However, this finding, particularly for anti-asthmatic medication usage, should be approached cautiously as it pertains to healthcare *use*. It may not fully represent the actual healthcare *needs* of the poorest children, who could be underrepresented due to limited access or utilization.

**Table 6:** Comparison of the least and the most affected by air pollution, depending on the outcome

|  | Average of the characteristics for : |  |  |  |  |  |
| --- | --- | --- | --- | --- | --- | --- |
|  | (i) the 10% the most affected,<br>(ii) the 50% the least affected, and (i)-(ii) difference most - least |  |  | (i) the 10% the most affected,<br>(ii) the 50% the least affected, and (i)-(ii) difference most - least |  |  |
|  | *Bronchiolitis emergency*<br>*before second anniversary* |  |  | *Antiasthmatic medication*<br>*before first anniversary* |  |  |
|  | (i) | (ii) | (i)-(ii) | (i) | (ii) | (i)-(ii) |
| Girl | 0.459 | 0.477 | -0.017(p=0.001) | 0.424 | 0.490 | -0.065(p=0.000) |
| Birth weight | 3094.33 | 3283.20 | -188.807(p=0.000) | 3229.00 | 3275.66 | -45.606(p=0.000) |
| Gestational age | 38.24 | 39.08 | -0.838(p=0.000) | 38.87 | 39.03 | -0.164(p=0.000) |
| No pathology | 0.693 | 0.831 | -0.136(p=0.000) | 0.800 | 0.821 | -0.020(p=0.000) |
| Cardio-respiratory pathology | 0.242 | 0.166 | 0.077(p=0.000) | 0.178 | 0.169 | 0.010(p=0.007) |
| Monitoring electrocardiogram | 0.116 | 0.039 | 0.076(p=0.000) | 0.060 | 0.043 | 0.017(p=0.000) |
| Premature birth | 0.187 | 0.059 | 0.127(p=0.000) | 0.095 | 0.069 | 0.026(p=0.000) |
| Low birth weight | 0.189 | 0.056 | 0.133(p=0.000) | 0.086 | 0.070 | 0.016(p=0.000) |
| Birth without significant issues | 0.581 | 0.741 | -0.160(p=0.000) | 0.689 | 0.731 | -0.043(p=0.000) |
| Disposable income percentile | 43.56 | 49.74 | -5.813(p=0.000) | 49.38 | 49.84 | -0.499(p=0.088) |
| Neonatology stay | 0.228 | 0.082 | 0.145(p=0.000) | 0.115 | 0.095 | 0.021(p=0.000) |
| Respiratory system radiography | 0.087 | 0.030 | 0.057(p=0.000) | 0.044 | 0.033 | 0.011(p=0.000) |
| Accessibility : GP | 3.97 | 4.10 | -0.127(p=0.000) | 4.04 | 4.10 | -0.051(p=0.000) |
| Accessibility : Paediatrician | 3.66 | 3.48 | 0.168(p=0.000) | 3.45 | 3.54 | -0.103(p=0.000) |
| Decile 1 | 0.174 | 0.090 | 0.083(p=0.000) | 0.111 | 0.094 | 0.018(p=0.000) |
| Decile 10 | 0.111 | 0.083 | 0.026(p=0.000) | 0.098 | 0.099 | -0.001(p=0.786) |
| CMU beneficiary | 0.171 | 0.133 | 0.036(p=0.000) | 0.121 | 0.130 | -0.010(p=0.005) |
| PM2.5 | 13.38 | 11.66 | 1.74(p=0.000) | 12.13 | 11.86 | 0.270(p=0.000) |
| Mother Age | 30.34 | 30.64 | -0.276(p=0.000) | 30.20 | 30.79 | -0.595(p=0.000) |
| Mother Born abroad | 0.238 | 0.164 | 0.074(p=0.000) | 0.167 | 0.166 | 0.003(p=0.390) |
| Outside Cities | 0.064 | 0.052 | 0.014(p=0.000) | 0.056 | 0.055 | -0.000(p=0.867) |
| Urban Area <50,000 | 0.094 | 0.112 | -0.018(p=0.000) | 0.112 | 0.108 | 0.004(p=0.128) |
| Urban Area 50,000 - 200,000 | 0.138 | 0.182 | -0.046(p=0.000) | 0.179 | 0.171 | 0.008(p=0.027) |
| Urban Area 200,000 - 700,000 | 0.176 | 0.231 | -0.054(p=0.000) | 0.203 | 0.225 | -0.023(p=0.000) |
| Urban Area >700,000 | 0.166 | 0.207 | -0.039(p=0.000) | 0.211 | 0.206 | 0.004(p=0.287) |
| Urban Area of Paris | 0.361 | 0.216 | 0.146(p=0.000) | 0.239 | 0.233 | 0.006(p=0.186) |
| Areas not EU-compliant | 0.794 | 0.729 | +0.066 (p=0.000) | 0.719 | 0.744 | -0.025 (p=0.000) |

Notes: (i) corresponds to group 4, with proxy predictor in the top 10% and group average treatment effect $\hat{\gamma}_4$ ; (ii) corresponds to group 1, with proxy predictor in the bottom 50% and group average treatment effect $\hat{\gamma}_1$ . Medians over 100 splits. Confidence intervals and p-values are as described in previous tables.

For any given health outcome, such as emergency admissions for respiratory issues or prescriptions for respiratory medications, it is pertinent to compare the group of children most affected by air pollution exposure with the broader cohort of all infants experiencing these health outcomes due to any cause. While air pollution exposure is a significant factor, it represents only one of many potential causes for such health issues in infants. This comparison helps to contextualize the specific impact of air pollution relative to other contributing factors. Figure 2 in the introduction provides a visual summary of our results. When comparing with the overall population distribution, we observe that the 8th decile of parental income suffers relatively the least, while the 1st decile suffers the most. Additionally, when comparing with the income distribution among children suffering from bronchiolitis due to all causes, we find that both ends of that distribution experience relatively more severe effects from bronchiolitis caused by air pollution compared to other causes. Specifically, the top decile in parental income accounts for 6.7% of all-cause emergency admissions for bronchiolitis, compared to 11.1% for admissions linked to air pollution. This result suggests that the top income decile may have a similar risk of seeing their children admitted to emergency for bronchiolitis due to air pollution as the rest of the population, while having a lower overall risk of being admitted to emergency for bronchiolitis due to all causes. One interpretation of this finding is related to their baseline, long-term exposure, potentially increasing their susceptibility to adverse effects from short-term pollution shocks.

Table 6 also shows that access to general practitioners (GP) and pediatricians is lower for those most affected, particularly regarding anti-asthma medication use, indicating potential accessibility challenges for more vulnerable groups. Concerning bronchiolitis, while access to a general practitioner is also lower for the most affected, access to a pediatrician appears to be relatively higher. This finding can be linked to the concentration of the most affected children in large urban areas, including Paris, where access to pediatricians is generally above average.

In an effort to better understand the role of health care accessibility, we incorporate variables measuring eligibility for CMU-C, an income-based complementary health insurance plan in France that eliminates all financial constraints on primary care for the poorest individuals. However, not all eligible individuals enroll, often due to lack of awareness or marginalization.

Our analysis, limited to 2010-2016 due to the need for fiscal income data, utilizes a CMU-C eligibility indicator based on household eligibility at birth or within two years thereafter, minimizing bias against low-income children.

We find that in the most affected group for anti-asthma medications, there is a higher likelihood of being in the bottom income decile, and a lower likelihood of unenrolled yet eligible CMU-C individuals, as Figure C1 shows. In contrast, for emergency bronchiolitis admissions, this group is more likely to be in the bottom income decile and eligible for CMU-C, but not necessarily enrolled. Given that these admissions are less avoidable, our results imply that observed healthcare consumption may underestimate the true effects, especially for non-emergency treatments, due to financial barriers limiting access to care. However, this bias is likely minor for emergency admissions.

Our results show that 79.4% of the most affected children (group 4) resided in areas where the annual PM2.5 exposure exceeded 10 *µg/m*^3^ in 2008, the European Commission’s 2030 target and the 2006 World Health Organization (WHO) guidelines. A higher baseline PM2.5 exposure correlates with increased sensitivity to air pollution for both analyzed outcomes, particularly bronchiolitis. Notably, the most affected group resides in municipalities where annual exposure levels are 1.7 *µg/m*^3^ higher than those least affected, measured in the year preceding birth to prevent the inclusion of endogenous variables as individual characteristics. This supports the interpretation that elevated baseline exposure intensifies the risk during short-term pollution spikes. We discuss in Appendix B an alternative explanation for this observation, which might stem from our study’s design: a thermal inversion may have a larger short-term impact on concentration levels in more polluted areas. We find supporting evidence for this hypothesis in one of the PM2.5 data sources, indicating that thermal inversions may have a more pronounced effect in highly polluted regions. Conversely, our other data source suggests a more uniform impact of the shock across different baseline exposure levels.

In summary, our results suggest that infants from the bottom 10% of parental income distribution face a higher risk of suffering the most from an air pollution shock. Conversely, the infants from the top 10% of parental income, who generally have a lower baseline risk, experience a relatively greater impact from air pollution compared to all other causes of bronchiolitis. Our research also provides evidence that factors such as healthcare accessibility and baseline exposure, which are influenced by the municipality of residence, may significantly contribute to these outcomes.

## 6 Policy implications

Effective air quality policies that yield tangible health benefits hinge on identifying and addressing the specific root causes of emissions and vulnerability factors unique to each area. As we detail below, doing so requires a comprehensive approach that includes data-driven analysis and tailored policy implementation. This section is devoted to the policy implications of our study, using the EU’s proposed revision of the Ambient Air Quality Directives as a framework for discussion.

This proposed policy package, aiming to align with the WHO’s guidelines by 2050, introduces interim air quality standards for 2030 and zero pollution targets by 2050, as part of a broader EU-wide climate neutrality objective. The framework includes three key elements which resonate with the findings of our study. First, it emphasizes the importance of public transparency, requiring improved air quality monitoring, reporting, and standardized indices across member states. Second, it is backed by a legal framework, including citizen compensation for air quality standard breaches. Lastly, the directives would require member states to create Air Quality Roadmaps to meet air quality standards, allowing for flexibility at the national and local levels to accommodate diverse economic and geographic conditions. This policy mirrors the framework of the Clean Air Act in the United States, where the State Implementation Plans enable states to tailor their own regulatory approaches to achieve and maintain national air quality standards. The 2008 Ambient Air Quality Directive already requires Member States to comply with pollution standards, based on local monitoring of concentrations. The French state was declared non-compliant with this directive by its highest administrative court in 2017 and has been fined 10 million euros every six months, recently reduced to 5 million euros.^32^

Our study underscores the need for enhanced air quality monitoring and harmonized indices in the EU, focusing on reducing measurement errors and providing detailed, accessible data for effective policymaking. Indices tailored to specific vulnerabilities, such as those of children and the elderly, could enhance the identification of “high-risk zones” for focused public health interventions and timely responses in areas most affected by pollution.

In addition, our findings also support the EU’s initiative to strengthen compensation rights for health damages from air pollution, particularly for vulnerable, often lower-income, populations. Emphasizing these groups in compensation strategies is important, considering their health care access challenges and financial constraints. Developing targeted compensation schemes could provide necessary financial relief, lessen health care burdens for these families, and promote adherence to air quality standards, functioning as both support and incentive to public and private entities.

More importantly, our results offer valuable insights for designing effective national air quality roadmaps. Our results suggest that these roadmaps should prioritize regions with vulnerable populations to mitigate severe health impacts of air pollution as early as possible. We illustrate the relevance of this targeted approach in our subsequent comparison of three different strategies for achieving the EU’s interim 2030 air quality targets for PM2.5 (annual average limit value of 10 *µg/m*^3^) in France.

### Air Quality Roadmaps: a tool for target interventions

Policies often target areas with high pollution levels. However, considering population vulnerability might offer a preferable strategy to maximize health benefits (Deryugina et al., 2021). Given the financial constraints associated with air pollution reduction, a priority ranking system that takes into account population characteristics could more effectively mitigate morbidity and mortality associated with exposure to air pollution. Such a targeted approach could accelerate the realization of benefits like reduced health care expenditures, fewer workdays lost to illness, and enhanced overall well-being, as compared to the conventional strategy of focusing solely on the most polluted areas.

From a practical standpoint, effective targeting necessitates identifying local PM2.5 emission sources, which may stem from vehicle emissions, industrial activities, energy consumption, agricultural practices, wildfires, or transboundary pollution. There is already a host mitigation policies available that, once local sources are identified, can be tailored accordingly. For example, measures to encourage the fast post-spreading burial of nitrogenous fertilizers and the use of covers for slurry pits in the agricultural sector are examples of targeted rural strategies to reduce emissions in the French National Plan for the Reduction of Atmospheric Pollutant Emissions for the period 2022-2025.^33^

We examine three distinct strategies for prioritizing the compliance of French municipalities with the 10 *µg/m*^3^ PM2.5 exposure threshold: one based on annual PM2.5 exposure levels, another on median income, and a third on the number of premature births. Over our study period, nearly 80% of most affected children were born in municipalities that did not meet the 10 *µg/m*^3^ exposure threshold. Assuming that the spatial distribution of the most vulnerable children across municipalities remains constant, our aim is to identify the most effective strategy for ensuring that these children reside in areas that comply with the exposure threshold as early as possible. Remark that targeting areas with a higher concentration of premature infants, not only addresses the immediate health impacts of PM2.5 exposure on these infants but also aligns with mitigating one of its possible root causes (Sun et al., 2015).

We employ two alternative methods for ranking municipalities. In the first approach, municipalities are grouped into small clusters that each represent 5% of all municipalities in France, regardless of the number of births. These clusters are formed based on similar values for either PM2.5 exposure, median income, or the number of premature births. Nonetheless, achieving the same reduction in a major city and in a smaller municipality may not be equally feasible. In the second approach, municipalities are grouped into clusters that account for 5% of total births nationwide, again based on similar values for the aforementioned variables. When we consider groups of municipalities that each account for an equal share of total births, achieving broad impact may require to prioritize a lower number of municipalities, but with greater population. For each clustering approach, the roll-out of compliance is then prioritized for clusters with either the highest levels of PM2.5 exposure, the lowest median income, or the greatest number of premature births.

Figure 10 provides the results when rolling-out compliance starting from the first group of municipalities (denoted p5) and extending to all (p100) municipalities that were non-compliant with the threshold in 2008. The results are presented for two different groupings: Figure 10a represents clusters that each account for 5% of all municipalities (irrespective of their size), while Figure 10b represents clusters that each account for 5% of total births nationwide (to account for size effects).

**Figure 10:**
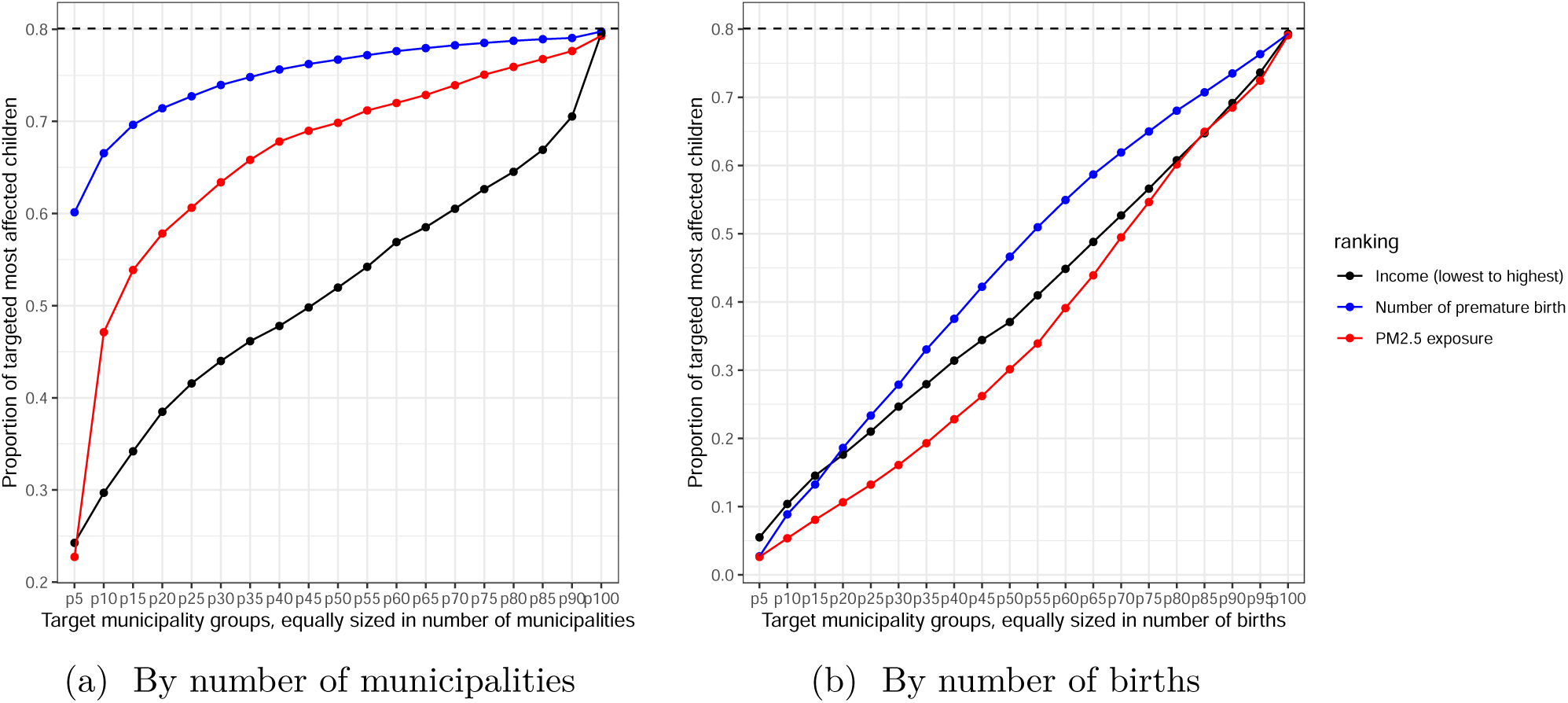
Comparative analysis of targeting strategies for achieving compliance with the 10 *µg/m*^3^ PM2.5 2030 target. *Notes: Non-compliant municipalities are ranked based on three criteria: PM2.5 exposure levels in 2008, incidence of premature births in 2018, and median income in 2018. Then, they are grouped into percentiles, either unweighted (left) or weighted by the number of births (right)*.

When clustering by municipalities, the most effective policy design appears to be targeting the top 5% of municipalities with the highest number of premature births, as this approach would immediately benefit 60% of the most vulnerable children. These municipalities account for nearly half (49.9%) of all births nationwide. Consequently, the financial burden of enforcing compliance in these areas could be substantial due to their large population. In contrast, focusing on the 10% of municipalities with the highest pollution levels (comprising 35.7% of all births) would reach 47% of the most affected children. Targeting based on median income proves to be the least effective approach in this case.

When clustering by number of births, the most effective approach remains to prioritize municipalities based on the number of premature births, provided that the target encompasses more than 20% of municipalities weighted by births. Targeting the lowest median income municipalities may be effective to achieve a short-term goal (maximize impact within the first 15% of municipalities weighted by births). Targeting municipalities with the highest PM2.5 exposure levels becomes the least effective strategy.

In both approaches, the conventional strategy that prioritizes the most polluted areas without considering local population characteristics is consistently dominated by at least one of the two alternatives. These alternatives, based on simple metrics, can be regarded as realistically implementable. Therefore, our study suggests that a more targeted approach, focusing on the most vulnerable populations, could more effectively deliver health benefits. Such an approach should inform the design of country-specific air quality roadmaps, ensuring that interventions are both effective and meaningful. It is important to note that our recommendations are primarily based on the effects of PM2.5 on young children and short-term health impacts; other pollutants, populations, and long-term effects are not considered, underscoring the need for further research and improved pollution data.

## 7 Conclusion

In this paper, we have examined the differential impacts of early childhood exposure to air pollution on children’s health care use across parental income groups using French administrative data. Our results provide quasi-experimental evidence linking vulnerability factors and pollution exposure on health measures during the first three years of life. In particular, we find causal evidence that short-term surges of air pollution affect the likelihood of emergency admissions and drug consumption related to respiratory issues for young children. We uncover substantial treatment effect heterogeneity using generic machine learning inference. Our analysis reveals that significant health effects of short-term variations of air pollution are concentrated in about 10% of the infant population. These infants are characterized by a combination of poor health indicators at birth, and are more likely to be from the lowest parental income decile. These effects should be seen as lower bounds, as they are measured using actual health care consumption, which only responds to adverse shocks when access is not an issue.

In light of our empirical findings, we advocate for a nuanced policy approach that goes beyond merely targeting regions with high pollution levels. Our analysis provides insights for the design of air quality policies, indicating that targeted interventions in regions with vulnerable populations may potentially lead to more immediate and substantial health benefits. However, it is important to carefully consider the composition of the population in these regions and evaluate the impact on all demographic groups to ensure a comprehensive and effective policy framework.

## Data Availability

All data produced in the present work are contained in the manuscript.

## Appendix A Data

### A.1 Hospital Discharge Data - PMSI

Our study relies on data from the PMSI MCO databases, which provide comprehensive information on each hospital admission in France, including inpatient stays and visits to emergency rooms. We identify hospital admissions for asthma or bronchiolitis that are associated with an emergency room visit using the main diagnoses codes (J45-J46 for asthma and J21 for bronchiolitis). These data accounted for patients whose entry mode was recorded as “home” and whose origin was “with emergency room visit”. However, these admissions do not include visits to the emergency room that did not result in hospitalization, or unplanned admissions with direct access to a hospital service other than the emergency department. Descriptive statistics of our main outcomes are provided in Table A2.

**Table A1:** Descriptive statistics on main outcomes (1/2)

| Birth year | By year |  |  | Through years |  | N |
| --- | --- | --- | --- | --- | --- | --- |
|  | First | Second | Third | First to Third anniversary | By Third anniversary |  |
| Drug delivery for obstructive airway diseases |  |  |  |  |  |  |
| 2008 | 0.238 | 0.217 | 0.184 | 0.296 | 0.393 | 35,831 |
| 2009 | 0.248 | 0.229 | 0.205 | 0.320 | 0.417 | 34,034 |
| 2010 | 0.244 | 0.246 | 0.202 | 0.328 | 0.420 | 33,892 |
| 2011 | 0.261 | 0.243 | 0.194 | 0.322 | 0.426 | 33,256 |
| 2012 | 0.270 | 0.247 | 0.208 | 0.333 | 0.437 | 34,910 |
| 2013 | 0.269 | 0.249 | 0.214 | 0.340 | 0.440 | 35,089 |
| 2014 | 0.266 | 0.252 | 0.206 | 0.338 | 0.439 | 34,658 |
| 2015 | 0.283 | 0.258 | 0.207 | 0.338 | 0.447 | 32,366 |
| 2016 | 0.278 | 0.255 |  |  |  | 31,254 |
| 2017 | 0.276 |  |  |  |  | 30,879 |
| All years | 0.263 | 0.244 | 0.202 | 0.327 | 0.427 | 336,169 |
| Visits to the Paediatrician |  |  |  |  |  |  |
| 2008 | 4.357 | 2.506 | 1.350 | 3.855 | 8.212 | 34,908 |
| 2009 | 4.437 | 2.515 | 1.355 | 3.869 | 8.307 | 33,441 |
| 2010 | 4.434 | 2.548 | 1.336 | 3.884 | 8.318 | 33,414 |
| 2011 | 4.441 | 2.517 | 1.302 | 3.818 | 8.259 | 32,845 |
| 2012 | 4.489 | 2.491 | 1.296 | 3.786 | 8.275 | 34,537 |
| 2013 | 4.600 | 2.428 | 1.254 | 3.682 | 8.282 | 34,769 |
| 2014 | 4.576 | 2.363 | 1.202 | 3.566 | 8.141 | 34,336 |
| 2015 | 4.494 | 2.279 | 1.168 | 3.447 | 7.941 | 32,101 |
| 2016 | 4.495 | 2.294 | 1.177 | 3.488 | 7.838 | 30,972 |
| 2017 | 4.581 | 2.348 |  |  |  | 30,518 |
| All years | 4.490 | 2.437 | 1.280 | 3.733 | 8.208 | 331,811 |
| Visits to the GP |  |  |  |  |  |  |
| 2008 | 7.265 | 6.520 | 5.201 | 11.721 | 18.986 | 34,908 |
| 2009 | 7.298 | 6.377 | 5.162 | 11.539 | 18.837 | 33,441 |
| 2010 | 7.182 | 6.341 | 5.003 | 11.343 | 18.526 | 33,414 |
| 2011 | 7.247 | 6.325 | 4.894 | 11.218 | 18.465 | 32,845 |
| 2012 | 7.252 | 6.219 | 4.815 | 11.034 | 18.286 | 34,537 |
| 2013 | 7.287 | 6.128 | 4.712 | 10.841 | 18.128 | 34,769 |
| 2014 | 7.201 | 6.040 | 4.490 | 10.530 | 17.730 | 34,336 |
| 2015 | 7.259 | 5.906 | 4.417 | 10.324 | 17.582 | 32,101 |
| 2016 | 7.185 | 5.807 | 4.344 | 10.239 | 17.334 | 30,972 |
| 2017 | 7.163 | 5.892 |  |  |  | 30,518 |
| All years | 7.235 | 6.182 | 4.825 | 11.049 | 18.294 | 331,811 |

**Table A2:** Descriptive statistics on main outcomes (2/2)

| Birth year | By year |  |  | Through years |  | N |
| --- | --- | --- | --- | --- | --- | --- |
|  | First | Second | Third | By Second anniversary | By Third anniversary |  |
| Emergency admission for asthma |  |  |  |  |  |  |
| 2009 | 0.004 | 0.005 | 0.004 | 0.009 | 0.012 | 34,034 |
| 2010 | 0.003 | 0.006 | 0.005 | 0.011 | 0.013 | 33,892 |
| 2011 | 0.004 | 0.007 | 0.005 | 0.012 | 0.015 | 33,256 |
| 2012 | 0.004 | 0.007 | 0.005 | 0.011 | 0.014 | 34,910 |
| 2013 | 0.004 | 0.006 | 0.006 | 0.012 | 0.015 | 35,089 |
| 2014 | 0.004 | 0.007 | 0.005 | 0.011 | 0.014 | 34,658 |
| 2015 | 0.005 | 0.008 | 0.006 | 0.013 | 0.017 | 32,366 |
| 2016 | 0.005 | 0.008 | 0.005 | 0.013 |  | 31,254 |
| 2017 | 0.006 |  |  |  |  | 30,879 |
| All years | 0.004 | 0.007 | 0.005 | 0.011 | 0.014 | 300,338 |
| Emergency admission for bronchiolitis |  |  |  |  |  |  |
| 2009 | 0.023 | 0.004 | 0.0005 | 0.026 | 0.027 | 34,034 |
| 2010 | 0.028 | 0.005 | 0.001 | 0.032 | 0.033 | 33,892 |
| 2011 | 0.032 | 0.005 | 0.001 | 0.036 | 0.037 | 33,256 |
| 2012 | 0.036 | 0.005 | 0.0005 | 0.040 | 0.040 | 34,910 |
| 2013 | 0.034 | 0.005 | 0.001 | 0.038 | 0.039 | 35,089 |
| 2014 | 0.031 | 0.005 | 0.001 | 0.036 | 0.036 | 34,658 |
| 2015 | 0.039 | 0.004 | 0.0005 | 0.043 | 0.043 | 32,366 |
| 2016 | 0.037 | 0.005 | 0.0003 | 0.041 |  | 31,254 |
| 2017 | 0.044 |  |  |  |  | 30,879 |
| All year | 0.034 | 0.005 | 0.001 | 0.036 | 0.036 | 300,338 |
| Emergency admission for bronchiolitis before second anniversary or asthma before third anniversary |  |  |  |  |  |  |
| 2009 | 0.026 | 0.009 | 0.005 |  | 0.036 | 34,034 |
| 2010 | 0.030 | 0.011 | 0.006 |  | 0.043 | 33,892 |
| 2011 | 0.035 | 0.012 | 0.006 |  | 0.048 | 33,256 |
| 2012 | 0.038 | 0.011 | 0.005 |  | 0.050 | 34,910 |
| 2013 | 0.036 | 0.011 | 0.007 |  | 0.051 | 35,089 |
| 2014 | 0.034 | 0.012 | 0.006 |  | 0.047 | 34,658 |
| 2015 | 0.043 | 0.012 | 0.006 |  | 0.056 | 32,366 |
| 2016 | 0.040 | 0.013 | 0.006 |  |  | 31,254 |
| 2017 | 0.047 |  |  |  |  | 30,879 |
| All year | 0.036 | 0.010 | 0.005 |  | 0.041 | 300,338 |

#### A.1.1 Birth hospital stay and baseline health indicators

Our data accounts for 99.6% of births in Metropolitan France (Quantin et al., 2013) , with the remaining 0.4% of births taking place outside of hospitals (Blondel et al., 2011). Newborn stays are identified in the PMSI using the criteria of the Scan Santé reference perinatal indicators. We also identify stays in neonatology units by the presence of a neonatology billing supplement (codes NN1, NN2, and NN3). We establish a general health status indicator based on principal diagnoses associated with the child’s stay. A child is considered healthy if its only primary diagnosis falls into codes Z38 “Children born alive, according to place of birth” or Z76.2 “Medical surveillance and care of other infants and children in good health”. The presence of a principal diagnosis associated with a condition, coded for example in chapter P “Certain conditions originating in the perinatal period”, is thus associated with at least one condition, without prejudging its severity.

A second indicator is proposed based on homogeneous groups of patients (GHM), a medico-economic nomenclature which makes it possible to describe and bill the health insurance system for the care of patients. This nomenclature evolved significantly in 2012 with regard to the major diagnostic category 15 “Newborns, premature babies and conditions of the perinatal period”, to be based firstly on age, then on the mode of entry, the presence of surgical procedures, the mode of discharge (in particular death) before looking at, by gestational age and weight group, the principal diagnosis with a closed list to detect “significant problems”. Newborns with “no significant problems” are classified as such by default, if there are no problems of greater severity that would have led to a different classification. In this approach, children are considered healthy at birth when they fall within codes 15M05A and 15M06A, i.e. “Newborns of 3300g and gestational age of 40SA and above with no significant problems” and “Newborns of 2400g and gestational age of 38SA and above with no significant problems”.

Furthermore, we introduced dummy variables based on the information from the child’s birth stay that indicate (i) whether any diagnosis pertains to respiratory or cardiovascular pathology codes, (ii) whether a radiography of the respiratory system was coded or not (PMSI MCO, Table A, if CDC-ACT starts with ZBQK00 or GEQH00 or LJQK00 or ZBQK003, encoded in Common classification of medical acts CCAM), and (iii) whether an electrocardiogram was reported (PMSI MCO, Table A, if CDC-ACT starts with “DEQP00”, in CCAM code).

#### A.1.2 Infant emergency hospital admissions by day of the week

We observe no significant seasonality in asthma emergency admissions on weekdays and weekends, whereas we see significantly fewer admissions on weekends for all respiratory admissions and bronchiolitis. (Table A3). This suggests that any episode of breathlessness in an infant with asthma or suspected asthma will lead to an emergency admission and is unavoidable regardless of the circumstances. This implies that the observed data is not biased by parental behavior or health care usage constraints. We obtain similar results for children less than 2 years old. For children less than 3 years old, there is evidence of fewer emergency admissions on weekends, in particular for the bottom 50% of parental income on Saturdays.

**Table A3:** Emergency admissions of EDP infants (less than one year old) by day of the week.

| Emergency admissions of EDP infants (less than one year old) - 2009-2017 |  |  |  |  |
| --- | --- | --- | --- | --- |
|  | All | J<br>Respi. | J21<br>Bronchiolitis | J45-J46<br>Asthma |
|  | (1) | (2) | (3) | (4) |
| Tuesday (ref: Monday) | 0.292<br>(0.750) | 0.475<br>(0.706) | 0.098<br>(0.523) | 0.292<br>(0.183) |
| Wednesday | -0.537<br>(0.750) | -0.235<br>(0.706) | -0.281<br>(0.523) | 0.156<br>(0.183) |
| Thursday | -1.096<br>(0.749) | -0.763<br>(0.706) | -0.159<br>(0.523) | -0.002<br>(0.183) |
| Friday | -0.740<br>(0.749) | -0.582<br>(0.706) | -0.366<br>(0.523) | -0.053<br>(0.183) |
| Saturday | -3.366***<br>(0.749) | -2.516***<br>(0.706) | -1.366***<br>(0.523) | -0.247<br>(0.183) |
| Sunday | -3.064***<br>(0.749) | -2.139***<br>(0.706) | -1.193**<br>(0.523) | -0.045<br>(0.183) |
| Constant | 15.932***<br>(0.530) | 13.414***<br>(0.499) | 6.872***<br>(0.370) | 2.134***<br>(0.130) |
| Observations | 3,287 | 3,287 | 3,287 | 3,287 |
| *Note:* |  | *p<0.1; **p<0.05; ***p<0.01 |  |  |

### A.2 Drug delivery data

We use data on drug dispensing for obstructive airway diseases from the DCIR database (*Données de Consommation Inter-Régimes*), which provides information on all reimbursed outpatient health care expenditures in France. A dispensing/delivery event corresponds to a pharmacy delivery medicine reimbursed by the health insurance fund for a medicine of class R03 “Drugs for obstructive airway diseases” of the Anatomical, Therapeutic, and Chemical (ATC) classification. The most common drugs are shown by parental income in Figure A1. The delivery of such medications is included in a reference algorithm for identifying chronic respiratory diseases. According to this criterion, the French National Health Insurance identifies individuals with chronic respiratory diseases as those who have received three dispensings of these drugs on separate occasions, except for those diagnosed with cystic fibrosis. Similar algorithms have been utilized in the medical literature, e.g. in Belhassen et al. (2016) and (Naiim et al., 2019), to identify cases of recurrent wheezing in infants and evaluate the short and long-term therapeutic management of asthmatic children, respectively. In a study combining the 2006 ESPS survey to health insurance reimbursement data, Delmas and Fuhrman (2012) demonstrated that one dispensing in 2005 and another in 2006 in ATC class R03 characterized 51.3% of self-reported asthmatics (both intermittent and persistent) aged 5 to 44 years, and those with such dispensings constituted 64.9% of self-reported asthmatics.

**Figure A1:**
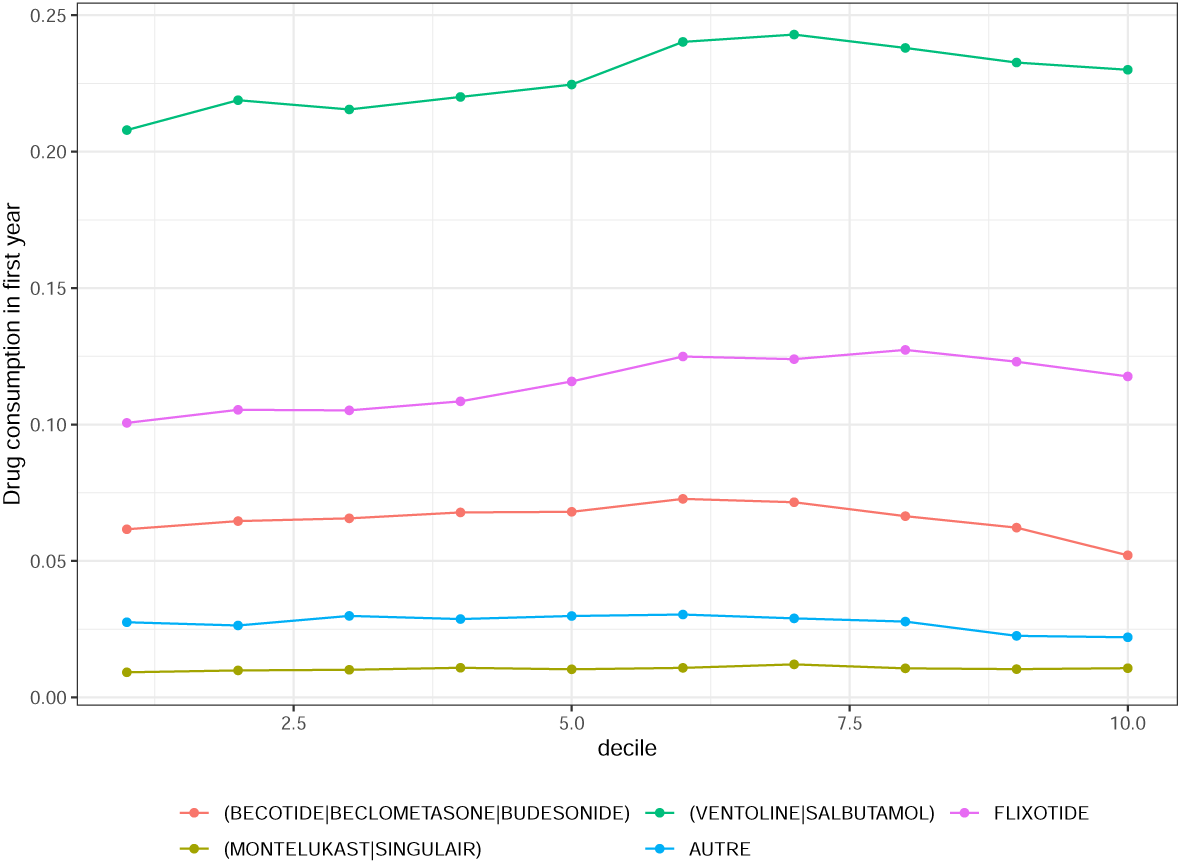
Drug deliveries to infants in their first year by income decile and main medications

### A.3 Air pollution data

Data on PM2.5 exposure is derived from two sources. The French National Institute for Industrial Environment and Risks (Ineris) produces historical data (2009-2020) as an annual average at the level of municipalities in metropolitan France (Ineris Cartothèque; Real et al. (2022)), based on a statistical comparison of observations from measurement stations and concentrations simulated by a numerical air quality model. Based on satellite observations, the Atmospheric Composition Analysis Group (Hammer et al., 2020) proposes annual average data at the European level on a one-kilometre grid (V4EU03, 2001-2018).

**Figure A2:**
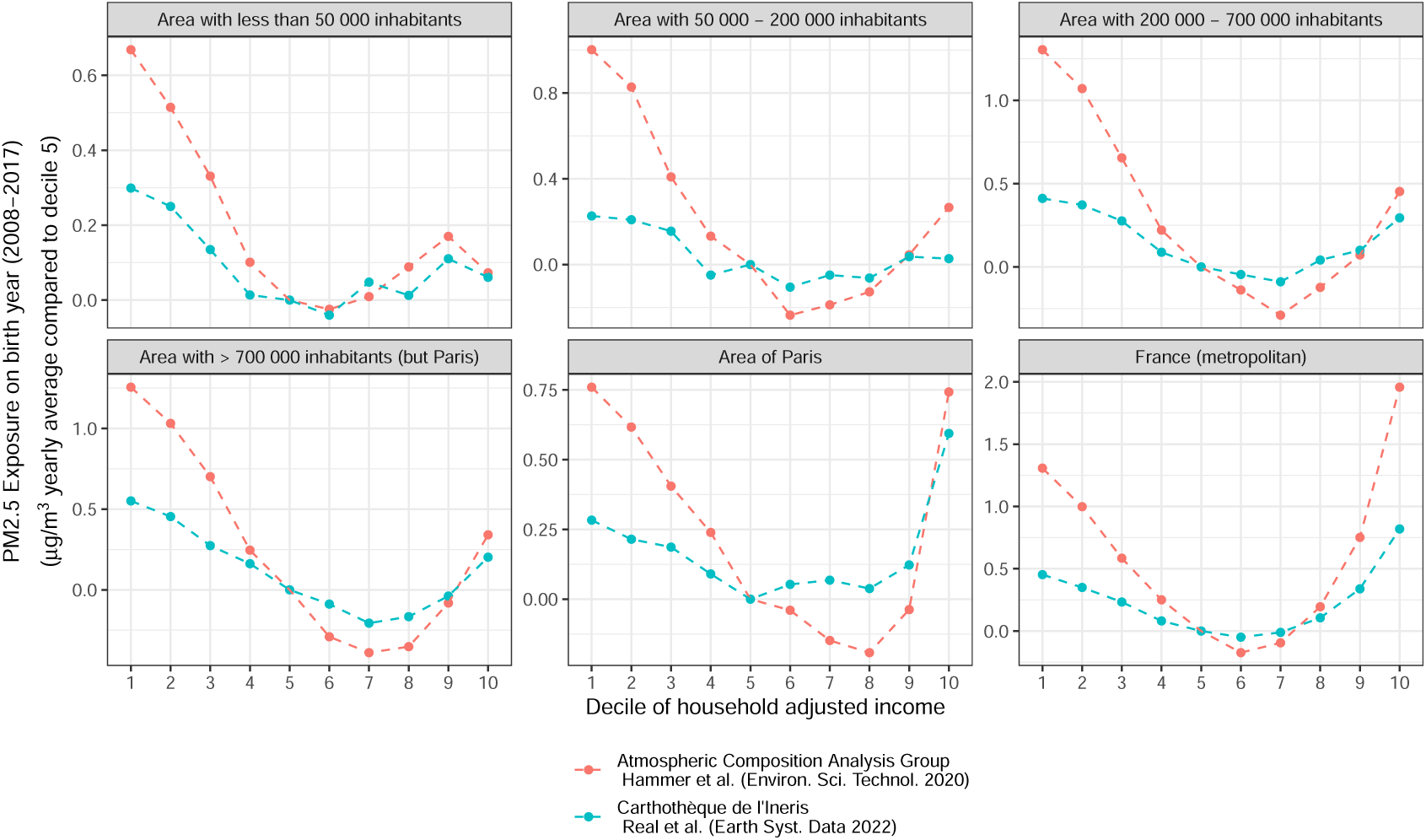
PM2.5 Exposure For Children in their Birth Year, By Urban Area, Relative to Fifth Decile of parental income

**Figure A3:**
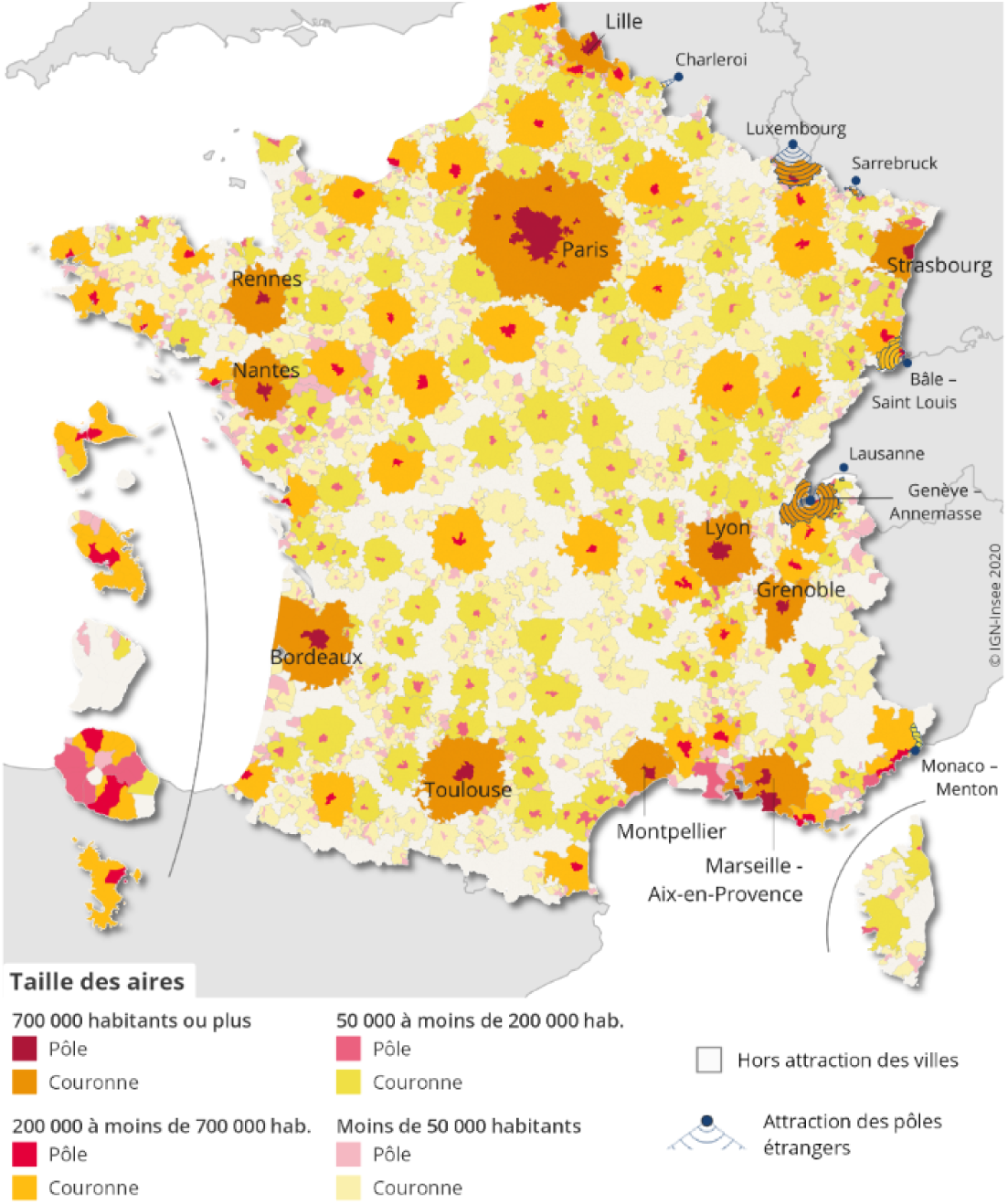
Urban areas by number of inhabitants in 2017 (Insee, 2020)

### A.4 Local weather data

The meteorological data, including the one used to identify days with thermal inversions, are those from the European atmosphere and surface reanalysis model (UERRA). This model generates hourly estimates of various meteorological variables such as temperature, humidity, and wind across multiple atmospheric layers, at a ground resolution of 11km x 11km. The UERRA project is laying the groundwork for a pan-European reanalysis with an extremely high resolution (5.5 km) forced by the global ERA-5 reference reanalysis system. As explained in UERRA’s user guide, atmospheric reanalysis is a technique used to reconstruct past weather conditions by combining historical observations (both in-situ and remote sensing via surface and satellite) with a dynamical model. The process generates a coherent description of the atmospheric state that is both physically and dynamically consistent. The model assimilates observational data to closely replicate the conditions it records. The main advantage of reanalysis is that it provides a multivariate, spatially complete, and coherent record of the atmospheric state, which is far more complete than any observational dataset.

These open-access data are available on the Copernicus website, in the Climate Data Store. We classify a day as a day with a thermal inversion when the difference between the daily average temperatures at 500 metres and 15 metres is positive. We count the number of such days over the first year of a child’s life. The meteorological control variables we use include the number of days in the year across 20 temperature intervals and 12 wind speed intervals (as per the Beaufort scale), as well as the annual average humidity and pressure.^34^

### A.5 EDP sample and linkage issues

The *Echantillon Démographique Permanent* (EDP) tracks individuals born on 16 specific birth dates (2nd to 5th of January; 1st to 4th of April, July, and October) across multiple survey and administrative sources. These sources are connected by a common identifier. Of the 364,105 EDP individuals with birth dates from 2008 to 2017, 340,897 have a recorded municipality of residence in metropolitan France. This information is issued from the birth certificate.

Out of these, 336,169 children are found in the National Health Insurance affiliates referential and consist of the main sample of our study. Approximately 98% had access to health care in their birth year. Given that France provides universal health insurance to its residents, the lack of access at this age likely suggests either absence from the territory or linkage issues.

### A.6 Adjusted income by birth cohort

We compute the adjusted disposable income per consumption unit at the household level for each infant. It is defined as the sum of earnings, self-employment income, capital income, and social transfers, minus direct taxes, and divided by the number of consumer units. The consumption units are computed using the modified OECD scale, which assigns 1 consumption unit to the first adult in the household, 0.5 to individuals aged 14 years or older, and 0.3 to children under the age of 14 years (Blanpain, 2019).

We rank infants within their birth cohort by the percentile of their household’s adjusted disposable income. We compute the corresponding percentile for each year from their birth year to two years later. 90% of infants in our primary sample born between 2008 and 2016 have at least one non-missing of these three percentiles. We define their income percentile as the average across the fiscal years in which the infant is observed. Restricting to the percentile in the birth year would result in attributing a percentile to only 66% of infants, forcing us to drastically limit our sample.

Moreover, among infants with at least one attributable percentile over their first three years, missing data about yearly adjusted income is correlated with being at the lower end of the income distribution, as depicted in Figure A4. If we limit the analysis to those infants for whom we only measure income in the birth year (y-axis of the Figure), we lose more children from low-income families than from high-income families (x-axis, when measured over up to 3 years). Specifically, this sample excludes all children born in 2017, as no measure of parental income is available for that year.

**Table A4:** Disposable parental income in 2016 Euros (precisely *Niveaux de vie*, equivalised disposable income in Eurostat terms)

| Decile | Interval | Median |
| --- | --- | --- |
| 1 | < 10,550 | 8,648 |
| 2 | 10,550 – 12,850 | 11,748 |
| 3 | 12,850 – 14,970 | 13,908 |
| 4 | 14,970 – 16,990 | 16,004 |
| 5 | 16,990 – 19,030 | 18,026 |
| 6 | 19,030 – 21,100 | 20,044 |
| 7 | 21,100 – 23,100 | 22,073 |
| 8 | 23,100 – 26,350 | 24,574 |
| 9 | 26,350 – 31,550 | 28,472 |
| 10 | > 31,550 | 38,384 |

**Figure A4:**
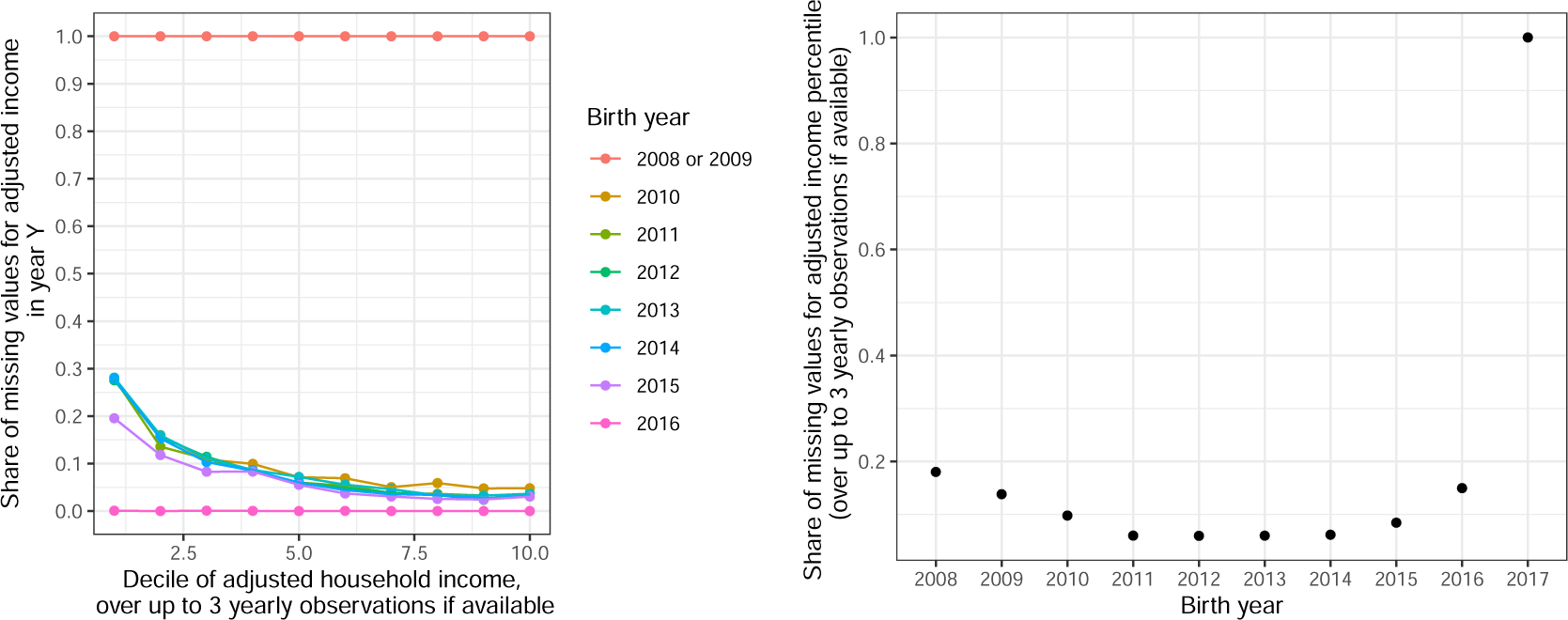
Household income missingness by birth year and decile of adjusted household income. Left: Share of missing values in birth year by final adjusted income decile. Right: Share of missing values in final adjusted income percentile (over up to 3 yearly observation if available in fiscal years 2010 to 2016).

To complete the descriptive statistics, Figure A5 shows the characteristics of childbirth stays by decile of adjusted household income for all non-premature births, and Table A5 reports the risk ratio between the bottom and the top of parental income distribution. This breakdown offers a comprehensive view of the socioeconomic factors influencing childbirth conditions and the care received. In addition, Figure A6 visualizes the frequency of emergency admissions for asthma and bronchiolitis by income decile, which reveal potential disparities in health outcomes or health care access among different socioeconomic groups.

**Table A5:** Risk Ratio (probability in the bottom tenth of parental income over probability in the top tenth of parental income)

|  | D1/D10 |
| --- | --- |
| Birth with a significant issue (GHM) | 1.04 [1.01;1.08] |
| Birth with at least one pathological diagnosis (DP des UM) | 1.08 [1.03;1.13] |
| Electrocardiogram at birth | 1.39 [1.23;1.56] |
| Neonathology at birth | 1.48 [1.35;1.60] |
| Respiratory radiography at birth | 1.75 [1.47;2.02] |
| Low birth weight (<2,5kg) | 1.77 [1.57;1.98] |

**Figure A5:**
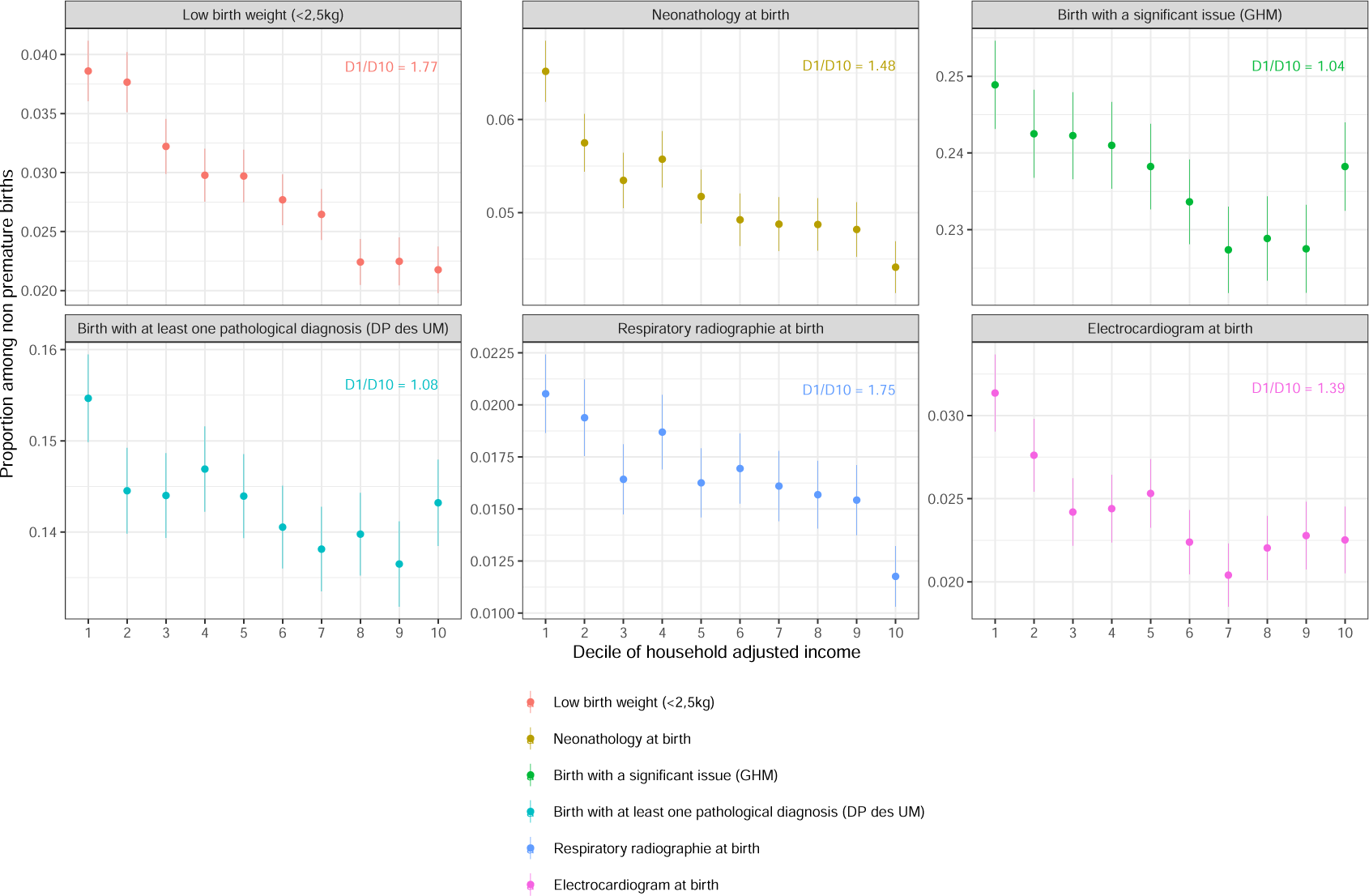
Characteristics of childbirth stays by adjusted household income among all non-premature births

**Figure A6:**
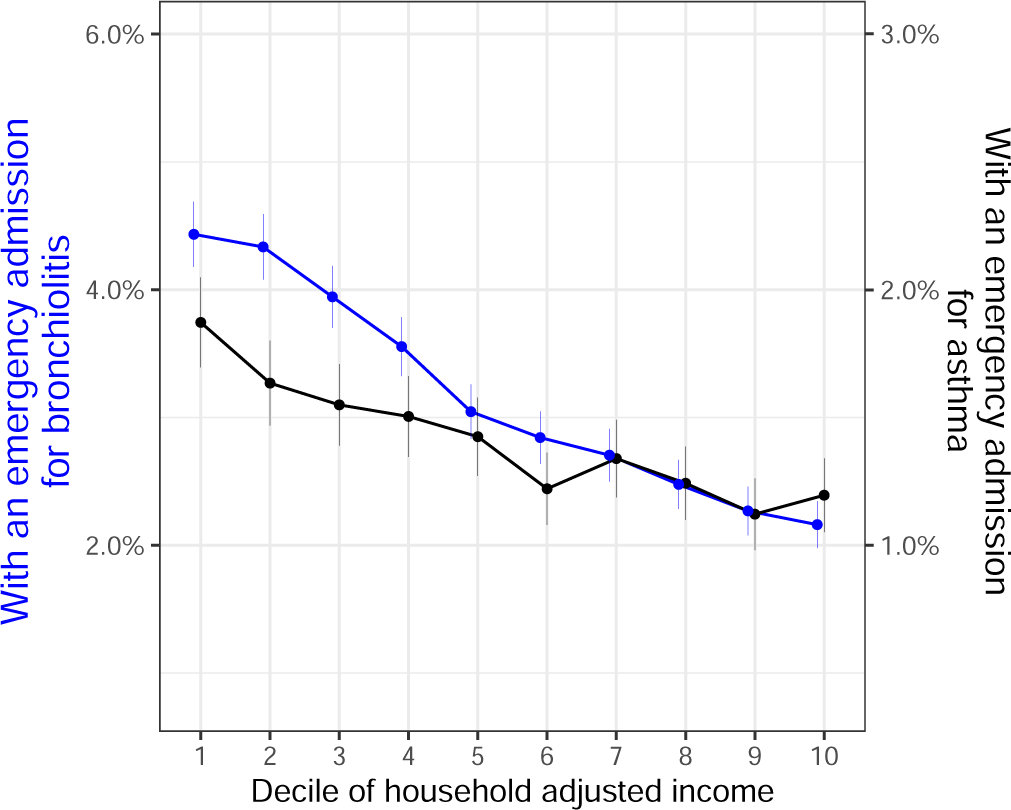
Respiratory Health indicators by adjusted household income

## Appendix B Treatment definition

### B.1 Thermal inversions

While thermal inversions have been used as instrumental variables in many studies due to their potential to generate as-good-as-random variations, they cannot be treated as completely random. Specifically, thermal inversion phenomena can be influenced by local topography, such as the presence of a deep valley, which can initiate or amplify the phenomena (Joly and Richard, 2018). Figure B1 illustrates how certain regions experience more thermal inversions than others over the years.

In our study, we aim to compare children born in the same municipality but at different times, thereby controlling for this local variation. We define the “usual” or “expected” number of days with thermal inversions by considering this long-term local average. This is achieved using the following linear model at the municipality and year level:

**Figure B1:**
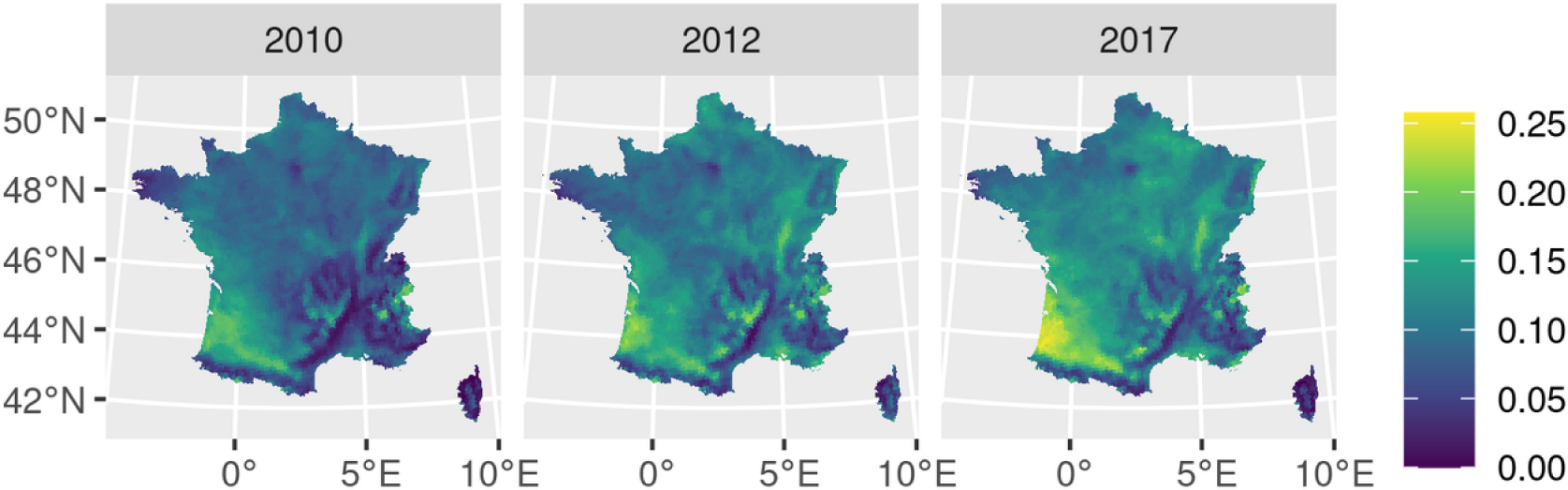
Share of days with thermal inversions for three example years

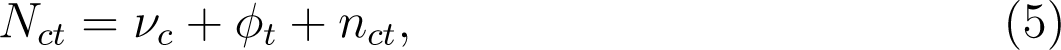

where *ν_c_* represents the long-term annual average of the number of days with thermal inversion, *ϕ_t_* is a national yearly shock, and *n_ct_* is a local idiosyncratic shock. According to its year of birth (*t*) and its municipality of residence (*c*) in its first year, each child (*i*) is assigned to the predicted local long-average *N̅_i_* = *N̂_ct_* = *ν̂_c_* + *ϕ̂_t_*. Figure B2 illustrates the geographical distribution of treated infants when using the adjusted number of days with thermal inversion, for two different thresholds. Compared to Figure B1, this map displays a more uniform distribution of treated cohorts than what we would have obtained if we had used the unadjusted number of days with thermal inversion to define the treated and untreated cohorts.

Thermal inversions typically follow a seasonal pattern, with a greater occurrence in winter. However, when these inversions are aggregated over the twelve months following a child’s birth, this seasonality tends to fade away as illustrated in Figure B3. Moreover, there are no clear trends based on the quarter of birth. On average, children born in January experience 36.4 days of thermal inversions in their first year, compared to 34.9 days for children born in April, 35.1 days for those born in July, and 35.2 days for those born in October. At most, there is a 1.5-day difference in total days of exposure to thermal inversion for children born in different quarters, which is significantly less than the threshold and size of the shock considered. However, one shortcoming of our approach is that we treat identically an additional 10 days of inversion happening in the first and the twelfth month of the child, whereas consequences may be quite distinct.

Table 2 shows that the characteristics of treated and non-treated children are remarkably similar. This similarity provides credibility to the assumption of exogeneity of our quasi-experimental shock, as it suggests that the exposure to thermal inversions is not systematically related to observable characteristics of the children.

**Figure B2:**
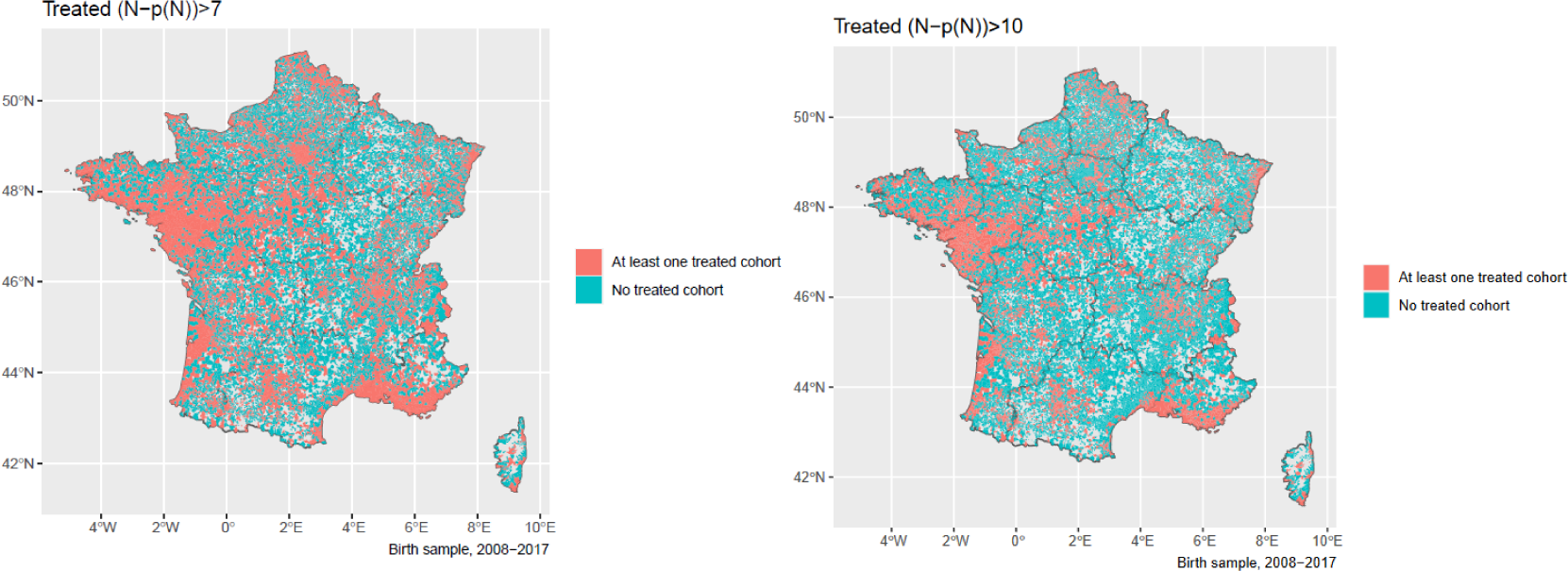
Municipality with at least one treated birth cohort over 2008 to 2017

**Figure B3:**
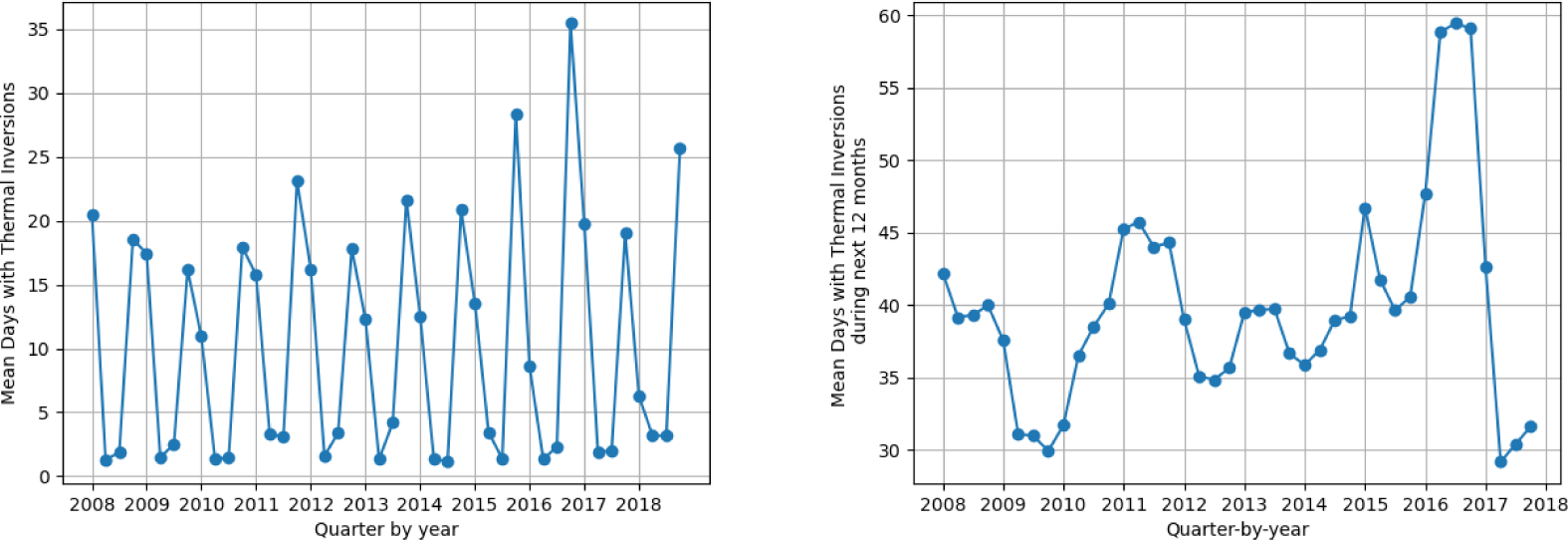
Temporal variations in the number of days with thermal inversions quarter by quarter, and in total the next 12 months

**Figure B4:**
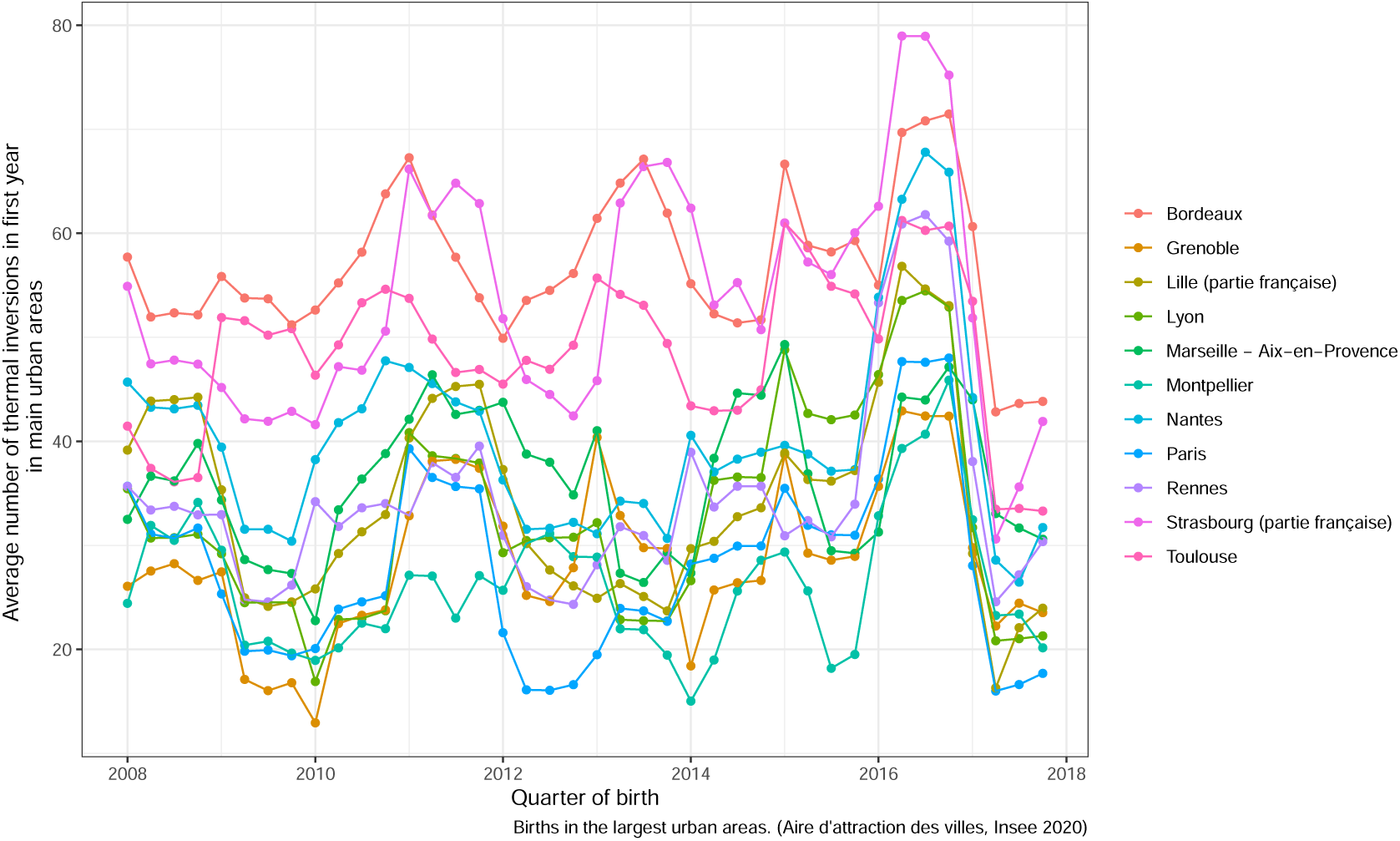
Temporal variations in the number of days with thermal inversions in children first years, by quarter of birth, in largest urban areas.

**Table B1:** Placebo treatment: average treatment effect of the quasi-experimental binary shock of exposure to air pollution after health care use

| | Intermediate exposure shock<br>$\bar{n} = 7$ | Large exposure shock<br>$\bar{n} = 10$ |
| --- | --- | --- |
| *Drug delivery for obstructive airway diseases* |  |  |
| - before first anniversary | 0.001<br>[-0.005;0.007] | 0.000<br>[-0.007;0.007] |
| Sample size | 288875 | 309758 |
| *Hospital emergency admissions* |  |  |
| - bronchiolitis or asthma before third anniversary | 0.001<br>[-0.002;0.004] | -0.001<br>[-0.005;0.002] |
| Sample size | 220155 | 233953 |
| - asthma before third anniversary | -0.0004<br>[-0.002;0.001] | -0.001<br>[-0.003;0.001] |
| Sample size | 220155 | 233953 |
| - bronchiolitis before second anniversary | 0.001<br>[-0.001;0.004] | 0.0002<br>[-0.003;0.003] |
| Sample size | 231904 | 251430 |

The estimates in Table 1 aligns well with daily findings presented by Godzinski and Suarez Castillo (2021), when considering an annual aggregation. According to their study, in days when thermal inversion occurs at each hour of the day, the mean concentration of PM2.5 rises by roughly 3 *µgm^−^*^3^. Additionally, other pollutants such as carbon monoxide and nitrogen dioxide also show significant increases. Therefore, if there is an extra 10 days with thermal inversion in a year, the annual mean concentration of PM2.5 would increase by approximately +(10× 3)/365 ≈ +0.1 *µgm^−^*^3^. We find slightly higher annual estimates, which may be explained by the fact that the daily estimates of the elasticity of PM2.5 to thermal inversion that we use for this back-of-the-envelope calculation are conditional on the inverse of planetary boundary layer height, an related instrument but different from to thermal inversions.

In Figure B5, we present a comparison of the exposure between the exposed group to the control group as a function of *n̄* which is equivalent to the data presented in columns (1) and (3) from Table 1. This data is broken down for both PM2.5 data sources, supplementing the information provided in Figure 7. Additionally, to supplement the information in Figure 8, we present in Figure B6 the data for emergency admissions, separately for bronchiolitis and asthma.

**Figure B5:**
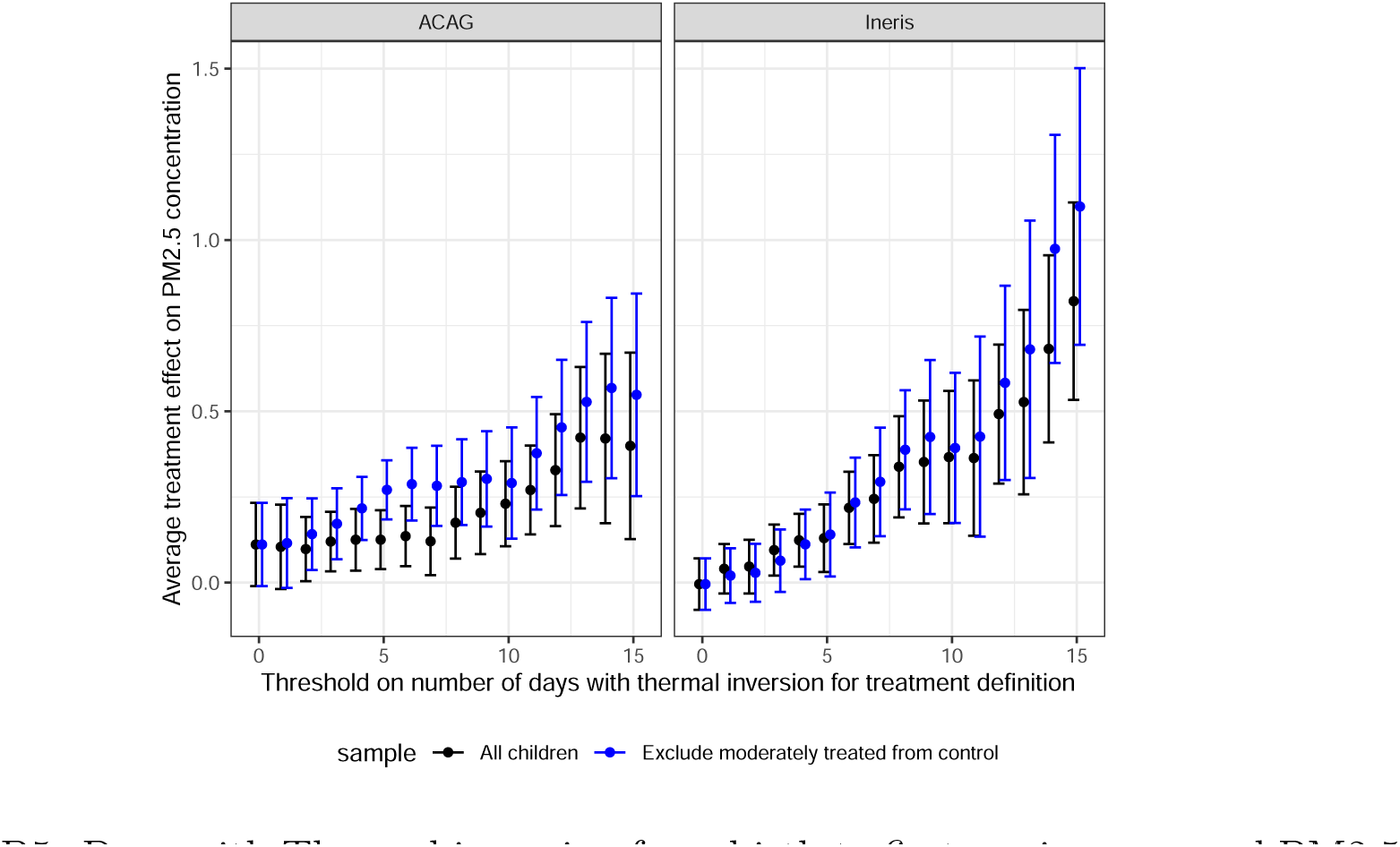
Days with Thermal inversion from birth to first anniversary and PM2.5 Exposure in first year. Notes: Infants born in January from the primary sample. Equation 3, coefficient *θ_n̄_*, excluding or not children with an intermediate level of treatment.

**Figure B6:**
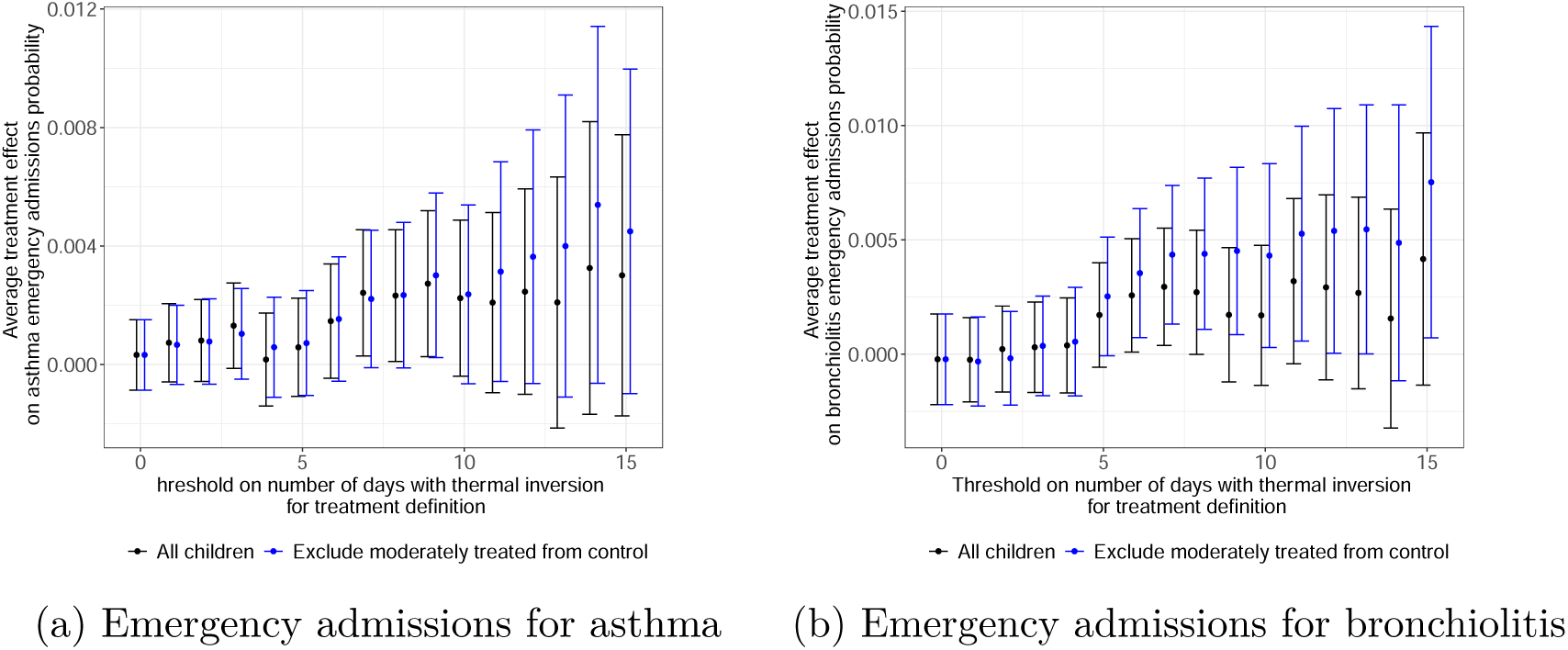
Days with Thermal inversion and Health Outcomes. Equation 2 (without *X_i_^I^*)

#### Baseline exposure levels

Thermal inversions may have a greater impact on PM2.5 concentrations in already polluted areas. Thus, one additional day with a thermal inversion could generate a greater increase in PM2.5 exposure, thus have stronger health impacts on those who live in large urban areas.

Our findings that the most affected children often reside in areas with higher baseline pollution levels can be interpreted in two ways: as a result of our instrumental variable approach, or as living in a highly polluted area being an additional risk factor for the effects of short-term air pollution fluctuations.

To explore the first channel, we modify (3) by including an interaction term with the treatment *T_i_*. This term considers whether, in the year prior to a child’s birth, their municipality had a PM2.5 concentration above the annual average, or if the child resides in an urban area with over 200,000 inhabitants. The outcomes of this analysis are detailed in Table B2. We find no heterogeneity by baseline exposure levels when considering the ACAG data source, but we do find some evidence of a greater impact of thermal inversions in overexposed areas with a significant interaction term of 0.453 (column (2) of Table B2).

**Table B2:**
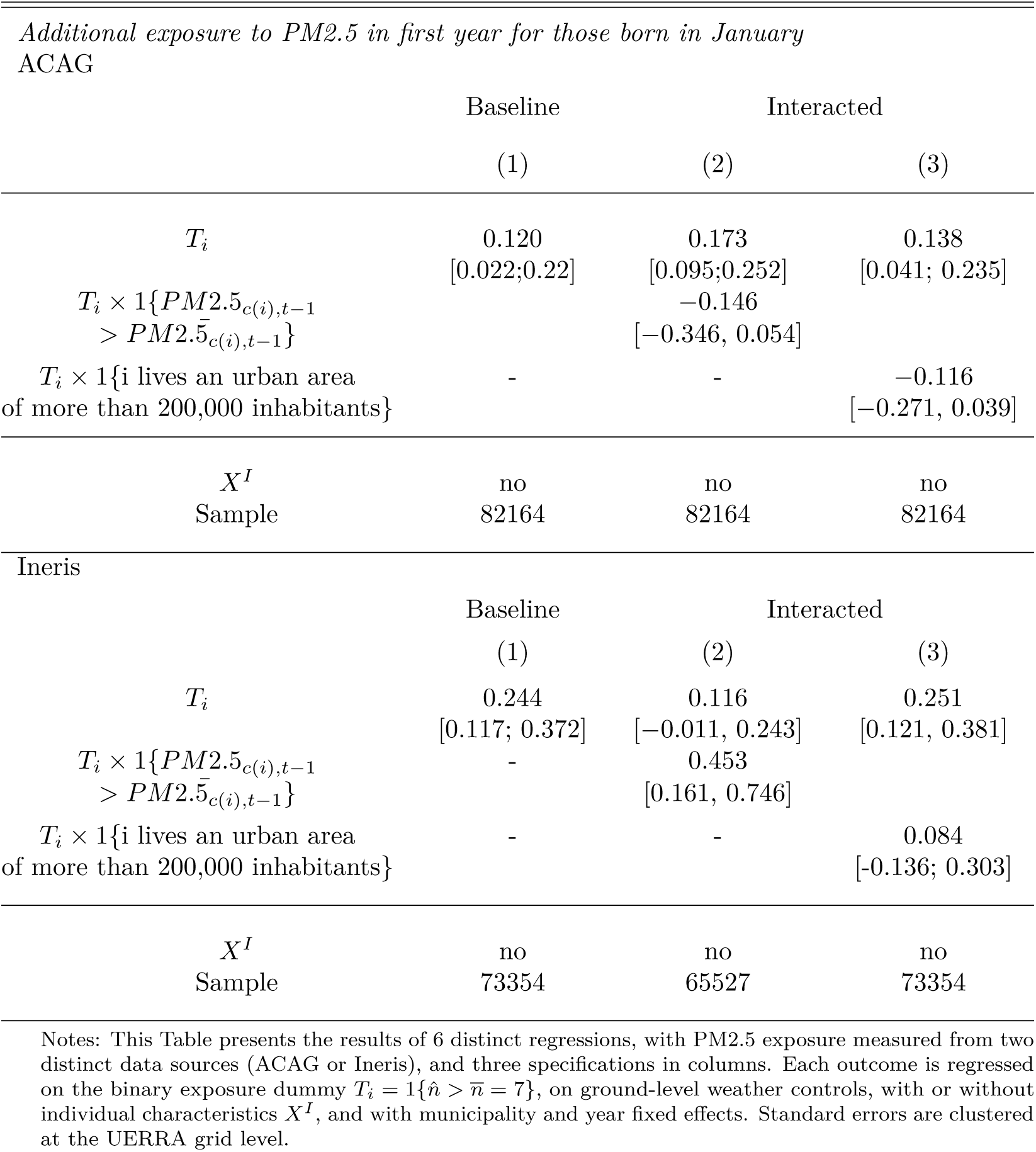
Design of the quasi-experimental treatment and exposure to air pollution depending on baseline pollution

### B.2 IV results

We present in this section the results of Instrumental Variable (IV) estimates using the quasi-experimental shock as an instrumental variable for PM2.5 annual exposure. We consider the equation (3) seen as a first stage with instrument *T_i_* for PM2.5 annual exposure of our two-sample two-stage instrumental variable estimator. We use a second-stage related to (2) to analyze a given health outcome as given by

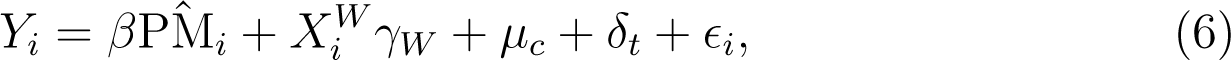

where PM̂*_i_* was obtained from the first-stage (3) estimated for a subsample, but we then extrapolate it to our full sample. Notably, we do not have observations of PM2.5 exposure before their first anniversary for the majority of children in our sample. We only have these observations for children born in January. This constraint necessitates a nonstandard approach to IV estimation (that is, not using the same sample for the first and the second stage) to maintain a fair sample size.

To be precise, we need to slightly modify the key identifying assumption stated in the main text, “after controlling for municipality fixed effects, year fixed effects, and weather variables, the local changes in the number of thermal inversions are unrelated to changes in the health outcomes of children *except through its influence on air pollution*” to “*except through its influence on PM2.5* ”.

In addition, we must explicitly assume that the estimate we obtain for the first stage (relationship between PM2.5 and thermal inversions) from the subsample of children born in January would not differ significantly from the corresponding estimates had we been able to run the estimation for all children. Given the well-documented relationship between PM2.5 and thermal inversion, we believe that the positive and significant relationship we found would still hold, though the exact magnitude could potentially vary. However, given the size of the confidence intervals obtained in the first stage, as shown in Table 1, and the consistency with estimates from Godzinski and Suarez Castillo (2021), it is likely that such estimates would fall within a similar range.

The two-sample two-stage instrumental variable estimation is described in Inoue and Solon (2010), and is asymptotically more efficient that a two-sample IV estimator. Zhao et al. (2019) emphasize the key assumption that the structural relations in the two samples should be the same: the exposure model should be correctly specified and in particular relies on the linearity assumption more heavily than in the one sample case. In our empirical application, the samples are not inherently different as they do not originate from separate populations; instead, the exposure model is estimated based on a subset. This approach offers certain advantages but also necessitates the above assumptions.

To perform inference, we take into account the geographically clustered nature of the data and of the treatment assignment process with a clustered pair-boostrap procedure. We first (1) form clustered bootstrap samples by resampling *X*^(^*^b^*^)^ = (*b*) (*b*) {*X*_1_ , · · · *, X_G_* } with replacement *B* = 500 times from the original sample *X* = {*X*_1_, · · · *, X_G_*}, where *X_g_* represents all the individual data attached to the *g*-th cluster, that is the UERRA grid unit (*G* = 4580), (2) compute on *X*^(^*^b^*^)^ the two-sample two-stage instrumental variable estimates *β̂* (3) compute the confidence interval at level at level *α* = 5% with the 2.5 and 97.5 percentiles of the estimates in step (2).

Table B3 reports the results, which are found to align with the corresponding reduced-form estimates presented in Table 3. The latter estimates correspond to the average treatment effects when scaled by a factor of 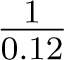 (source: ACAG) or 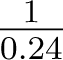 (source: Ineris).^35^ In our primary analysis, these average treatment effects were assessed as the consequence of an increase in PM2.5 annual exposure by 0.12 to 0.24 *µg/m*^3^, along with potential impacts of other pollutants, resulting from approximately 10 days of substantial increase in air pollution exposure during the child’s first year of life. The IV estimates, while bearing the caveats discussed earlier, can be directly interpreted as the causal effect of a 1 *µg/m*^3^ increase in PM2.5 annual exposure during the first year.

**Table B3:** IV estimates with endogenous variables PM2.5 (annual mean exposure). 500 bootstraps of the two-sample two-stage instrumental variable estimation.

| *Outcome* | **Source: ACAG**<br>(1) | **Source: Ineris**<br>(2) |
| --- | --- | --- |
| *Having drug delivery for obstructive airway diseases before first anniversary* | 0.048 [-0.017;0.167] | 0.023 [-0.005; 0.053] |
| *Hospital emergency admissions for bronchiolitis or asthma before third anniversary* | 0.057 [0.006;0.207] | 0.024 [0.005; 0.062] |
| *Hospital emergency admissions: asthma before third anniversary* | 0.026 [0.002;0.083] | 0.011 [0.002;0.027] |
| *Hospital emergency admissions: bronchiolitis before second anniversary* | 0.028 [0.002;0.11] | 0.014 [0.001;0.038] |

### B.3 Controlling for quarter of birth

The incidence of respiratory diseases depends on infant quarter-of-birth because quarter-of-birth is related to the age at the epidemic season onset, a risk factor well-known for bronchiolitis (Fauroux et al., 2020). In our sample, infants born in October are at significant higher risk of bronchiolitis emergency admission than infant born in April, which are older than six months when the winter season occurs, as shown in Figure B7. We first note no discernible seasonal pattern in the annual count of thermal inversions children experience based on their quarter-of-birth, a factor which could have indicated an incidental confounding factor. Nonetheless, it is essential to explore how incorporating the quarter-of-birth variable might influence our overall analysis.

We first define a new long-term average as the “expected” quarterly number of days with thermal inversions from *N_cyq_* = *ν_c_* + *ϕ_y_* + *α_q_* + *n_cyq_*, and assign to each child *i* resident of municipality *c* a prediction of the number of “expected” days with thermal inversions *N̅_i_* as the sum of *N̂_cqt_* over the four first quarters *y, q* of child *i*. Except for this adjustment to take into account quarterly dynamics in the prediction, exposed and unexposed cohorts are defined as before (13.4% are in the treated group against 14.1% in our baseline model), and we obtain very close results to Table 1 (relationship between PM2.5 and being in the exposed cohort) and Table 3 (relationship between respiratory diseases outcomes and being in the exposed cohort) without additional adjustment.

**Figure B7:**
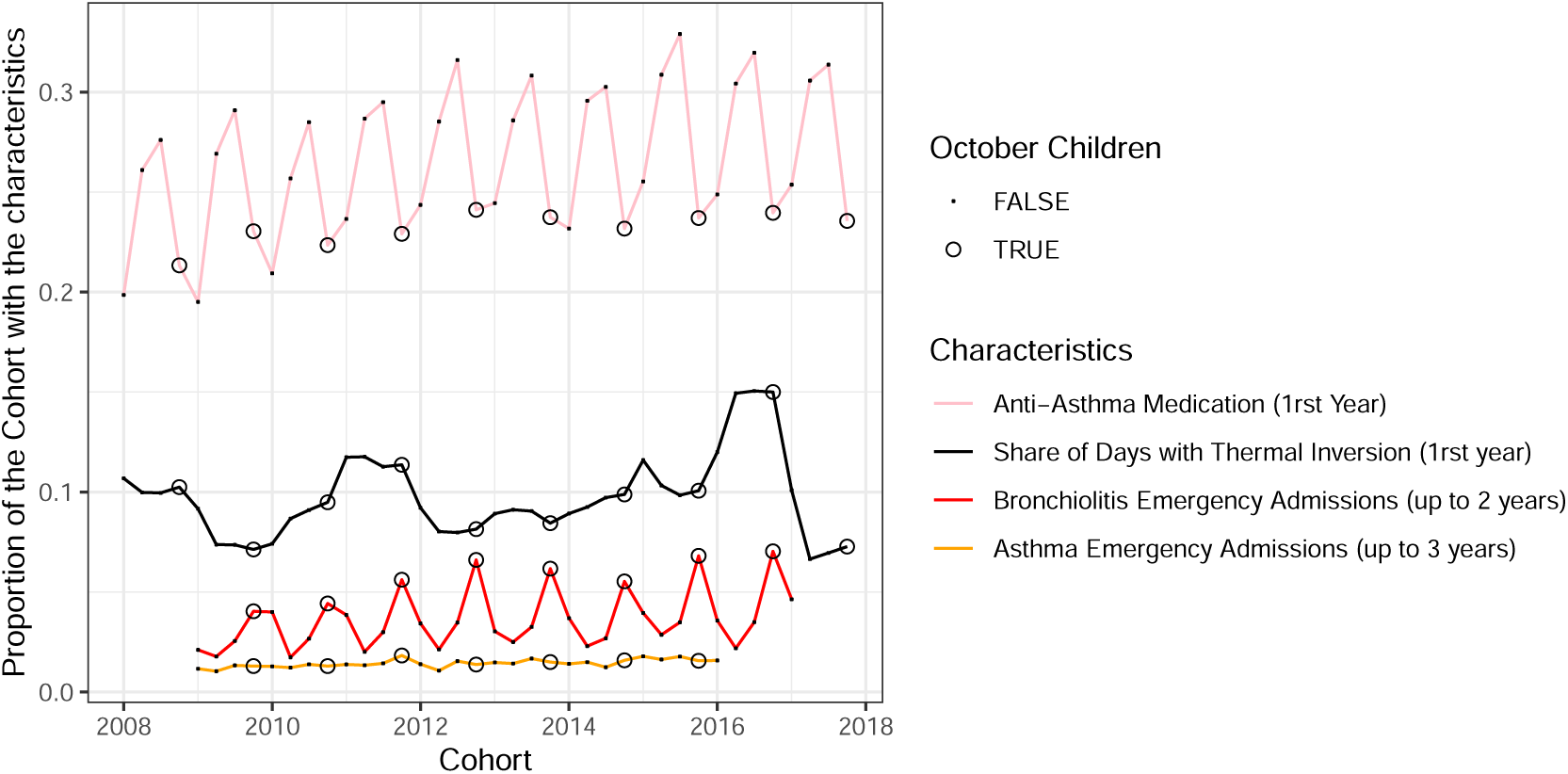
Seasonality of Outcomes and Exposure

In Table B4, we show how our main set of results are modified when controlling for quarter-of-births. As for emergency admission for bronchiolitis, if anything, the quarter model in column (3) and (4) points to more precise estimates. In line with the absence of quarter-of-birth seasonality in emergency admissions for asthma, the point estimates are very close but precision slightly drops. Finally, the point estimate for anti-asthma medication, already non-significant in the baseline, remains non-significant and changes sign. To obtain a positive and significant estimates, whereas in the baseline specification, a threshold of 10 additional days with thermal inversion was enough, in this case, one has to choose a threshold higher than 15 days (point estimates of 0.009 [-0.002; 0.019], p-value = 0.10).

All-in-all, opting for a quarterly model has little to no implications for our results concerning emergency admissions for respiratory diseases, but render results for anti-asthma medication more fragile.

**Table B4:** Average treatment effect of the quasi-experimental air pollution shock

| *Outcome* | Baseline |  | Quarter-of-Birth Model |  |
| --- | --- | --- | --- | --- |
|  | (1) | (2) | (3) | (4) |
| *Having drug delivery for obstructive airway diseases before first anniversary* |  |  |  |  |
|  | 0.003 | 0.003 | -0.004 | -0.007 |
|  | [-0.003;0.009] | [-0.004;0.010] | [-0.010;0.002] | [-0.014;0.000] |
| Quarter-of-birth control | no | no | yes | yes |
| $X^I$ | no | yes | no | yes |
| Sample size | 336169 | 217859 | 3336169 | 217859 |
| *Hospital emergency admissions: bronchiolitis or asthma before third anniversary* |  |  |  |  |
|  | 0.005 | 0.007 | 0.004 | 0.004 |
|  | [0.002;0.009] | [0.003;0.011] | [-0.000;0.007] | [0.000;0.008] |
| Quarter-of-birth control | no | no | yes | yes |
| $X^I$ | no | yes | no | yes |
| Sample size | 246075 | 189722 | 246075 | 189722 |
| *Hospital emergency admissions: asthma before third anniversary* |  |  |  |  |
|  | 0.002 | 0.003 | 0.002 | 0.002 |
|  | [0.000;0.005] | [0.001;0.006] | [-0.000;0.004] | [-0.001;0.005] |
| Quarter-of-birth control | no | no | yes | yes |
| $X^I$ | no | yes | no | yes |
| Sample size | 246075 | 189722 | 246075 | 189722 |
| *Hospital emergency admissions: bronchiolitis before second anniversary* |  |  |  |  |
|  | 0.003 | 0.003 | 0.003 | 0.004 |
|  | [0.000;0.006] | [-0.000;0.006] | [0.001;0.006] | [0.001;0.007] |
| Quarter-of-birth control | no | no | yes | yes |
| $X^I$ | no | yes | no | yes |
| Sample size | 277271 | 207825 | 277271 | 207825 |

*Notes:* This table presents the results of 16 distinct regressions, with 4 outcomes in rows and 4 specifications in columns. Each outcome is regressed on the binary treatment designed to capture a positive variation in air pollution exposure, $T_i = 1\{\hat{n} > \bar{n} = 7\}$ , quantifying whether a child was over-exposed to thermal inversion in its first year, on ground-level weather controls, with or without individual characteristics $X^I$ , municipality and year fixed effects. In columns (3) and (4), a control for quarter-of-birth is introduced. Standard errors are clustered at the UERRA grid level.

### B.4 Another outcome: doctor visits

Finally, we can consider another outcome that is not specific to respiratory diseases but is related to children’s use of health care: doctor visits. We consider all doctor visits, and GP and paediatrician separately and present the results in Table B5. For total doctor visits, we find a modest yet significant increase that remains consistent across all four specifications. An increase of 0.1 to an average of 11.7 visits represents an increase of about 1%, which appears to be mainly driven by an increase in visits to paediatricians, i.e. specialists, a finding consistent with the need to first see a doctor who can diagnose and prescribe anti-asthmatic drugs. In unreported results, we find evidence of heterogeneity in the BLP sense, but do not succeed in separating groups of impact in the GATEs sense.

**Table B5:** Doctor’s visits: Average treatment effect of the quasi-experimental air pollution shock

| *Outcome* | All Children |  | Excluding those intermediately treated |  |
| --- | --- | --- | --- | --- |
|  | (1) | (2) | (3) | (4) |
| *Visits to GP or Paediatrician before first anniversary* |  |  |  |  |
|  | 0.10 | 0.099 | 0.14 | 0.12 |
|  | [0.022;0.18] | [0.014;0.18] | [0.041;0.25] | [-0.002;0.23] |
| $X^I$ | no | yes | no | yes |
| Sample size | 331811 | 216814 | 232331 | 146337 |
| *Visits to Paediatrician before first anniversary* |  |  |  |  |
|  | 0.064 | 0.064 | 0.082 | 0.060 |
|  | [0.001;0.13] | [-0.011;0.14] | [0.008;0.16] | [-0.037;0.16] |
| $X^I$ | no | yes | no | yes |
| Sample size | 331811 | 216814 | 232331 | 146337 |
| *Visits to GP before first anniversary* |  |  |  |  |
|  | 0.037 | 0.035 | 0.062 | 0.056 |
|  | [-0.050;0.12] | [-0.062;0.13] | [-0.038;0.16] | [-0.063;0.18] |
| $X^I$ | no | yes | no | yes |
| Sample size | 331811 | 216814 | 232331 | 146337 |

*Notes:* This table presents the results of 12 distinct regressions, with 3 outcomes in rows and 4 specifications in columns. Each outcome is regressed on the binary treatment designed to capture a positive variation in air pollution exposure, $T_i = 1\{\hat{n} > \bar{n} = 7\}$ , quantifying whether a child was over-exposed to thermal inversion in its first year, on ground-level weather controls, with or without individual characteristics $X^I$ , municipality and year fixed effects. Standard errors are clustered at the UERRA grid level.

## Appendix C Generic machine learning estimation of heterogenous treatment effect

In this section, we discuss the methodology introduced by Chernozhukov et al. (2023) and its application in our context. The method aims to: (1) detect any observable heterogeneity in treatment effects; (2) determine the specific treatment effects across different impact groups; and (3) identify covariates linked to this heterogeneity. Given a large array of potential covariates influencing vulnerability to treatment (Table C1), testing each for heterogeneity risks overfitting. Conversely, limiting the analysis to a pre-selected subset of variables could omit crucial insights. This approach systematically utilizes all available covariates to identify meaningful heterogeneity. Its strength lies in its applicability to any machine learning algorithm and its provision of a valid inference procedure.

**Table C1:** List of Z features

|  |  |
| --- | --- |
| Socio-economic charac. | Gender |
|  | Mother age |
|  | Mother born abroad (indicator) |
|  | Decile (indicators) and percentile (linearly) of parental income |
|  | Benefit from the CMU-C |
| Baseline health<br>(from birth hospital stay) | Birth weight (indicator of low birth and linearly) |
|  | Gestational age (indicator of being premature and linearly) |
|  | Healthy birth |
|  | Normal Birth |
|  | Stay in the neonatology department |
|  | Respiratory or cardiovascular diagnosis |
|  | Electrocardiogram at birth |
|  | Radiography of the respiratory system at birth |
| Local charac. | PM2.5 exposure (the year before birth) |
|  | Accessibility to GP |
|  | Accessibility to paediatrician |
|  | Type/size of the urban area (6 modalities) |

Each of the objectives (1), (2) and (3) is associated with estimates based on a proxy predictor *Ŝ*(*z*) of the conditional average treatment effect *s*_0_(*Z*) and were named as follows: (1) the best linear predictor of the conditional average treatment effect (BLP), (2) the sorted group average treatment effects or the average treatment effects by group induced by the proxy (GATES) and (3) the classification analysis or the average characteristics of the least and most affected units (CLANS).

We aim to characterize the conditional average treatment effect function, *Z* → *s*_0_(*Z*) = E[*Y* |*Z, T* = 1] − E[*Y* |*Z, T* = 0]: which set of *Z*’s characterizes those who are the most affected? if not all children are affected, what proportion of the children population has a significant average treatment effect? The basic step requires to randomly divide the sample in two, an auxiliary sample *A* and a main sample *M* . Sample *A* is used to estimate a proxy (non necessarily consistent) of *s*_0_, *z* → *S_A_*(*z*), and while using sample *M* , *S_A_* can be treated as a fixed map and applied to each infant to obtain a vulnerability index, to later confirm (or not) its relevance in term of heterogeneity of the treatment effects, in the same way we could use with a pre-defined covariate : statistical inference is then conceptually straightforward. However, repeating sample split requires to appeal to quantile-aggregated inference – which aggregates inferential results by taking medians of estimates and medians of upper and lower confidence intervals obtained from different splits.

### C.1 Estimation

Let us denote each observation by *i* (infant). Our numerical implementation makes use of *S* = 100 random splits of the data. Over each random split, we define a main sample M and an auxiliary sample A over each half of the sample split. We train and tune two ML algorithms (random forest and lasso) to predict: (1) the conditional average treatment effect *s*_0_(*Z_i_*) = E[*Y_i_*|*Z_i_, T_i_* = 1]−E[*Y_i_*|*Z_i_, T_i_* = 0] and, (2) the outcome for infants in the control group *b*_0_(*Z_i_*) = E[*Y_i_*|*Z_i_, T_i_* = 0], using sample A: (*Y_i_, T_i_, Z_i_*)*_i∈A_*. We proceed by separately estimating E[*Y_i_*|*Z_i_, T_i_* = 1] and E[*Y_i_*|*Z_i_, T_i_* = 0], an approach known as “T-learners”. An alternative approach would be to use the Horvitz-Thompson transformation, multiplying the outcome by the standardized treatment 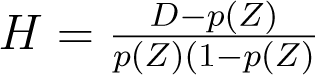, so that the expectation of *HY* is the conditional average treatment effect itselft, which can directly be fitted in one step, in an approach known as “causal regression”.

We thus obtain two proxies for these functions of interest (for each ML method): *z* → *S_A_*(*z*) and *z* → *B_A_*(*z*). We use them to estimate the best linear predictor (BLP), the group-average treatment effects (GATES), and perform a classification analysis (CLAN) as follows.

#### BLP

For each main sample, we estimate the best linear predictor by weighted OLS, with weights {*p̂*(*X_i_*)(1 − *p̂*(*X_i_*))}*^−^*^1^, from the equation

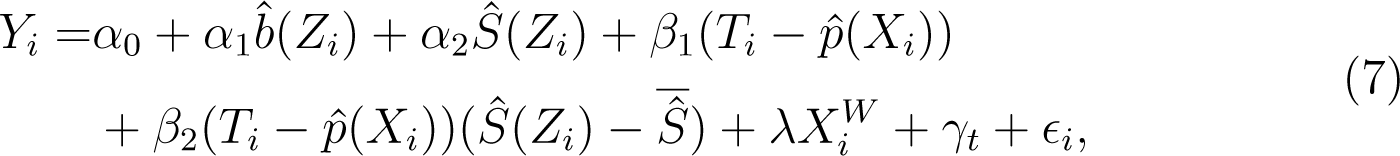

where *Z_i_* combines individual-level variables *X_i_^I^* and exposure variables, such as PM exposure, GP and pediatrician accessibility, and size of urban areas. We also control for *X^W^* surface-level weather variables and for year-of-birth fixed effects.

The best linear predictor of *s*_0_(*z*) is *β̂*_1_ + *β̂*_2_*Ŝ*(*z*). It provides information about treatment effect heterogeneity. Specifically, *β̂*_2_ is a consistent estimate for the correlation between the sensibility to treatment, as measured by the proxy predictor, and the true conditional average treatment effect, *Cov*(*s*_0_(*Z*)*, S*(*Z*))*/V ar*(*S*(*Z*)). Testing the null hypothesis *β*_2_ = 0 hence provides a way to investigate the presence of heterogeneous treatment effects, i.e. that *s*_0_ varies with *Z*, and the relevance of *S*(*Z*), i.e. that it is correlated with *s*_0_. In practice, we may include any “noise-reducing” proxy function of *Z* in this equation. In practical experiment, Chernozhukov et al. (2023) indicate that including *B̂* significantly improves the precision of estimation of the BLP. In the last version of their working paper, they also mention including *p̂*(*Z*), and its interaction with *Ŝ*(*Z*). In further investigation, we could test whether in our case, such modifications improve or not precision.

#### GATES

For each main sample, we create four groups *k* = 1, 2, 3, 4 of increasing predicted average treatment effects based on the 50th, 75th, and 90th quantiles of *Ŝ*(*Z_i_*). We use these groups to estimate the group-average treatment effects by weighted OLS, with weights {*p̂*(*X_i_*)(1 − *p̂*(*X_i_*))}*^−^*^1^, from

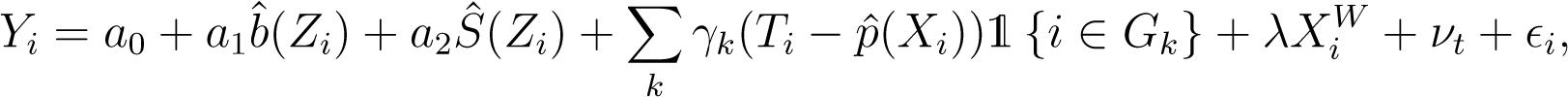

where *γ̂_k_* denotes the expectation of *s*_0_ in group k, and *G_k_*’s denote group membership. By construction of *Ŝ*(*Z_i_*), the GATES should be such that *γ̂*_1_ ≤ *γ̂*_2_ ≤ *γ̂*_3_ ≤ *γ̂*_4_ since these parameters are interpreted as the respective average treatment effects of each group of sensibility to the treatment. This approach delivers valuable information about the distribution of the negative health impacts of the quasi-experimental pollution shock across children in our data. In absence of heterogeneity or if the proxy fails to capture it, it should be that all the GATES are statistically the same. Here as well, we may include any “noise-reducing” proxy function of *Z* in this equation. In the last version of their working paper, they also mention including *p̂*(*Z*), and its interaction with 1{*i* ∈ *G_k_*}.

#### CLAN

Finally, for each main sample, we investigate the compositions of the *G_k_* groups defined in the GATES using the vector of CLAN parameters

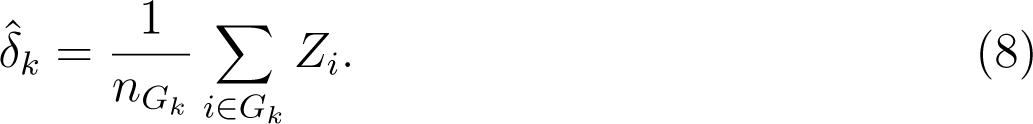

These estimates correspond to the average characteristics of each group *G_k_* along the multiple dimensions of *Z*. They hence allow drawing a portrait of the different groups, in particular the most affected children in *G*_4_ for which *Ŝ*(*Z_i_*) exceeds its 90th quantile.

### C.2 Inference

Conditionally on a single split, the estimate *θ̂* ∈ (*β̂, γ̂, δ̂*) is approximately Gaussian, as the sample size *N* , leading to a partition (⌊*N/*2⌋, ⌊*N/*2⌋), goes to infinity following standard statistical inference.

The median aggregated p-values across split to test whether one of these estimates is distinct from zero is defined as the minimum between *p*^+^, the median across split 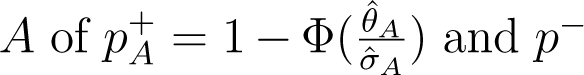 and *p^−^*, the median across split 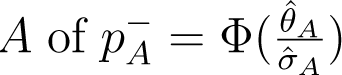. Under weak assumption, the probability that this p-value, multiplied by two, is below the level *α* is close to *α* when (⌊*N/*2⌋, ⌊*N/*2⌋) goes to infinity, that is the standard property. In addition, under the median concentration condition, the factor 2 is not necessary. This condition states that the approximate median over-the-splits t-statistics 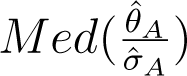 tend to concentrate more than any single-_̂_ *σ̂_A_* split t-statistic 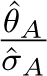, a condition that Chernozhukov et al. (2023) find mild and *A* supported by their computational experiments. Although they do not recommend multiplying by 2, we do so in this version for presenting p-values, which may be rather conservative. P-values should be divided by two otherwise.

Median confidence intervals behave similarly: if the final level of coverage *α* is required, without the median concentration condition, one should aggregate by taking the median the lower and upper bound confidence interval at level *α/*2. With the median concentration condition, the factor 2 is not necessary. We present confidence intervals which may be read as 90% confidence intervals under the conservative view or 95% confidence intervals under the median concentration assumption.

We report their median across the splits, and their confidence intervals of coverage 1 − *α* = 95%, as the median across the splits of CIs with coverage 1 − *α/*2. We adapt the numerical routines from the GenericML R package (Welz et al., 2022) to allow for propensity scores outside of [0.05,0.95], as well as random sample splits with up to 95% of control units (90% in source codes). In our implementation, we compare the performance of the lasso and random forest to form the two proxy predictors.

The performance metrics are based on the two empirical counterpart of respectively 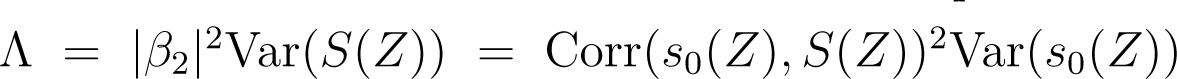, and Λ^̄^ = 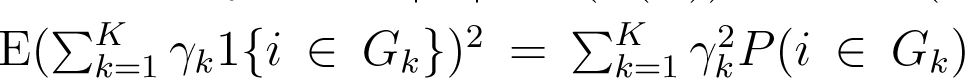. The first is used to choose the best algorithm for the BLP analysis as the maximizer of the correlation between the proxy *S*(*Z*) and the CATE *s*_0_(*Z*) to find heterogeneity. The second is used to choose the best algorithm for the GATES and CLAN analysis, as the maximizer of the *R*^2^ of the regression of *s*_0_(*Z*) on 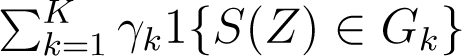. We report these in Table C2.

**Table C2:** ML performance metrics

| Best BLP ($\Lambda$)<br>$\vert \beta_2\vert ^2 \text{Var}(S(Z))$ | | Best Gates ($\bar{\Lambda}$)<br>$\sum_{k=1}^K \gamma_k^2 P(i \in G_k)$ | |
| --- | --- | --- | --- |
| Lasso | Random Forest | Lasso | Random Forest |
| *Having drug delivery for obstructive airway diseases before first anniversary* |  |  |  |
| 3.216 | 114.101 | 43.077 | 141.480 |
| *Number of drug deliveries for obstructive airway diseases before first anniversary* |  |  |  |
| 7.609 | 16.762 | 26.822 | 28.315 |
| *Visits to GP or Paediatrician before first anniversary* |  |  |  |
| 6.269 | 4.497 | 2.620 | 3.230 |
| *Hospital emergency admissions for bronchiolitis or asthma before third anniversary* |  |  |  |
| 2.680 | 39.627 | 34.438 | 40.815 |
| *Hospital emergency admissions for asthma before third anniversary* |  |  |  |
| 10.725 | 18.785 | 11.377 | 10.938 |
| *Hospital emergency admissions for bronchiolitis before second anniversary* |  |  |  |
| 1.706 | 179.189 | 10.779 | 34.385 |

Median over 100 splits, multiplied by $10^6$ . For the BLP, the chosen algorithm is the one that reaches the highest estimated $\Lambda$ . For the GATES and the CLAN, the chosen algorithm is the one that reaches the highest estimated $\bar{\Lambda}$ .

#### Access to healthcare: CMU-C

Figure C1 provides the CLANs focused on variables measuring accessibility to healthcare.

**Figure C1:**
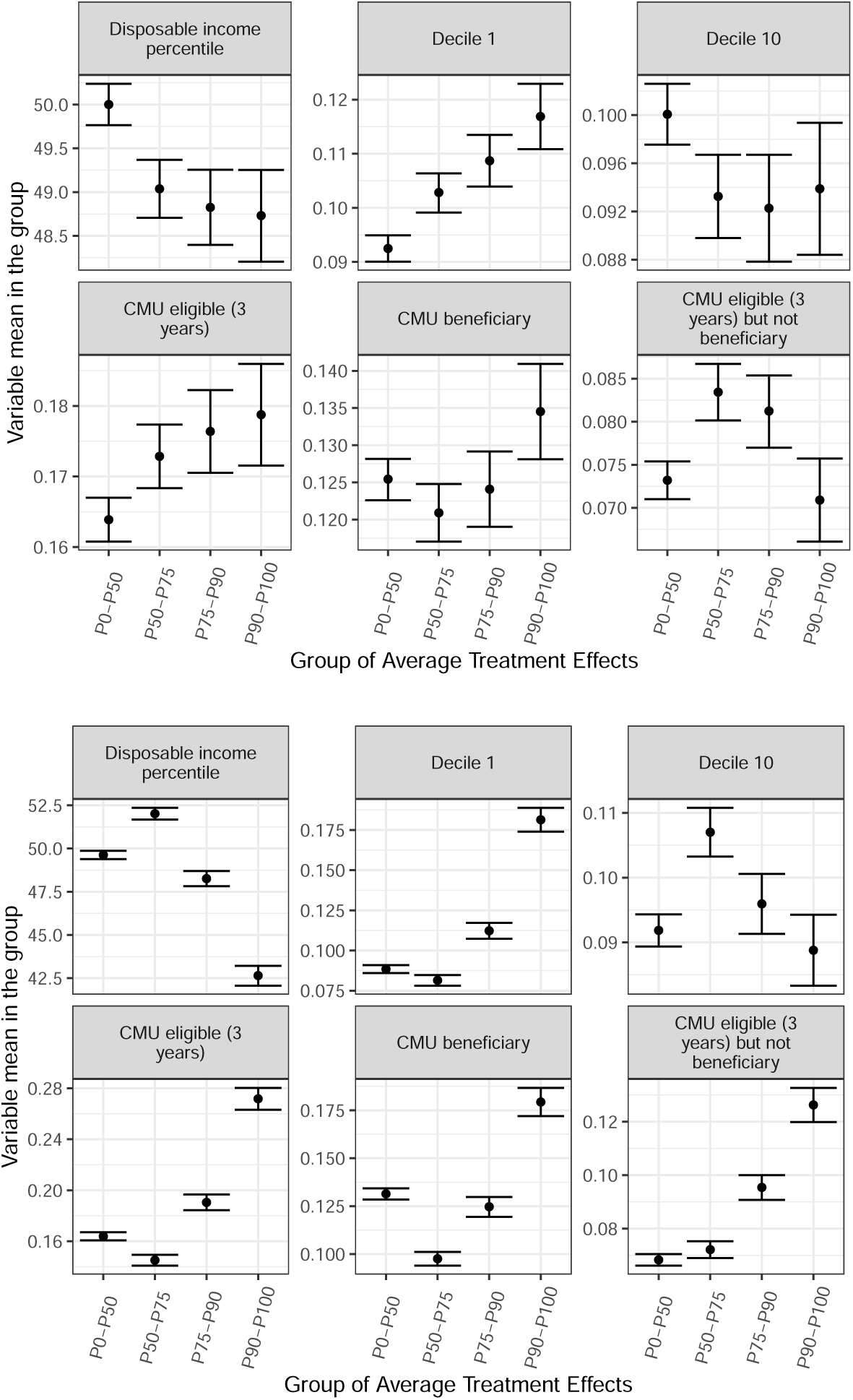
CLANs of CMU-C variables. Top: Obstructive Airways Diseases - Drug Consumption. Bottom: Emergency admission for bronchiolitis.

## Footnotes

1 Medications for asthma address both immediate symptoms and chronic management. Deschenes, Greenstone and Shapiro (2017) emphasize that these drugs also act as a defensive investment by reducing asthma-related hospitalizations and fatalities despite the condition’s persistent prevalence (Fanta, 2009).

2 According to Open medic (CNAM), this drug class represents 9.9% of the euro value of drug deliveries reimbursed for individuals under 20 years old. The average cost amounts to 58.5 euros per young beneficiary (under age 20) in 2017, for an average of 3.7 packs, of which 41.3 euros are reimbursed by the National Health Insurance.

3 Amendments adopted by the European Parliament on 13 September 2023 on the proposal for a directive of the European Parliament and of the Council on ambient air quality and cleaner air for Europe (recast) (COM(2022)0542 – C9-0364/2022 – 2022/0347(COD))(1).

4 Additional information is provided in Appendix A.

5 These instructions were given to the parents through the “Carnet de Santé”, a notebook given at birth and filled in by health professionals throughout the child’s development.

6 “Bilan de la qualité de l’air extérieur en France en 2017.” Rapport Commissariat Général au Développement Durable, 2018, Ministère de la Transition Ecologique et Solidaire.

7 “Bilan de la qualité de l’air 2019, Surveillance et information en Ile-de-France.”, 2019, Air Parif.

8 We are grateful to the Copernicus Climate Change and Atmosphere Monitoring Services for making the Copernicus products available. We acknowledge that neither the European Commission nor ECMWF assumes responsibility for any use that may be made of the Copernicus information or data it contains.

9 The median area of the municipality regions (approximately 11 square kilometers) aligns, on average, with the dimensions of the UERRA grid units.

10 Jans, Johansson and Nilsso^n^ (2018) and Dechezleprêtre, Rivers and Stadler (2019) employ this definition for thermal inversions as well.

11 Specifically, these dates are the 2nd, 3rd, 4th, and 5th of January, and the 1st through 4th of April, July, and October.

12 Conducted by the Direction de la Recherche, des Études, de l’Évaluation et des Statistiques

13 Commonly prescribed molecules include salbutamol and fluticasone propionate; details in Appendix A.

14 Appendix A.1.2 shows evidence of the unavoidability of asthma admissions, indicating that emergency admissions correlate with actual infant health and not differential healthcare access.

15 https://www.santepubliquefrance.fr, 2023, 9th of April.

16 Haute Autorité de Santé, 2009, “Asthme de l’enfant de moins de 36 mois : diagnostic, prise en charge et traitement en dehors des épisodes aigus.” *Recommandations Professionnelles*

17 As defined by the French Federation of Perinatal Health Networks (ATIH) and the DREES in ScanSanté/Indicateurs de santé périnatale/FFRSP/ATIH.

18 Details on income groups and consumer unit calculations are in Table A4 in the Appendix, using the modified OECD scale.

19 To address this, researchers commonly control for time-invariant and unobserved local determinants using location fixed effects.

20 To tackle this issue, some studies use time variations that are credibly unrelated to residential sorting or transient economic factors (Isen, Rossin-Slater and Walker, 2017; Jans, Johansson and Nilsson, 2018). For instance, they compare cohorts born before or after the introduction of significant environmental regulations, like the 1970 Clean Air Act, or investigate short-term health responses to specific events such as thermal inversions.

21 Moreover, weather-related IVs impact pollutants beyond PM2.5, complicating the identification of effects attributable to a specific pollutant. This complexity challenges the interpretation of 2SLS estimates, as demonstrated by Godzinski and Suarez Castillo (2021).

22 Appendix B.2 provides the results of a two-sample two-stage least squares regression, where the first-stage is given by (3) and the second-stage corresponds to (2), using *PM_i_* instead of *T_i_*.

23 These health indicators include (i) neonatalogy department stay, (ii) respiratory or cardiovascular diagnoses, (iii) electrocardiogram, (iv) respiratory system, (v) no pathology diagnosis, (vi) a birth labeled as “normal” for national insurance reimbursement purposes.

24 These estimates align with daily estimates from Godzinski and Suarez Castillo (2021) when aggregated yearly (see Appendix B.1).

25 The impact of excluding intermediately treated children appears to be more pronounced with ACAG data, but it is also noticeable with Ineris data, particularly for larger threshold values *n*, as shown by Figure B5 in the Appendix.

26 Comparing births based on extreme PM2.5 exposure levels (e.g., the 10% most exposed versus the 50% least exposed) would not be suitable primarily because of the spatial sorting driven by household characteristics, which could introduce biases.

27 The propensity score *p*(*X_i_^I^*) is specified as a logit model, where *X_i_^I^* includes the same individual variables used in (2).

28 For clarity, we have omitted a key estimation step for consistency of (*β̂*_1_*, β̂*_2_): the treatment needs to be residualized with the propensity scores, and the OLS needs to be weighted with the Horvitz-Thompson transform of propensity scores (Chernozhukov et al., 2023).

29 We adapt the numerical routines from the GenericML R package (Welz et al., 2022) to allow for propensity scores outside of [0.05,0.95], as well as random sample splits with up to 95% of control units (90% in source codes). In our implementation, we compare the performance of the lasso and random forest to form the two proxy predictors.

30 In Appendix B.4, we further examine the number of doctor visits as an additional outcome, although non-specific to respiratory diseases, and find a modest yet significant increase that remains consistent across all four specifications.

31 In comparison, Deryugina et al. (2019) report a lower correlation, between 0.013 and 0.015, for wind-induced exposure’s impact on elderly mortality. This may be attributed to the higher difficulty of predicting individual-level daily mortality risk, given the rarity of the event.

32 This fine reduction decision acknowledges air-quality improvement plans in non-compliant areas like Paris and Marseilles. Local initiatives include the *Plan de protection de l’Atmosphère*, that detail local measures such as with low emission zones or portside electricity for boats. Nationally, actions encompass funding for air quality efforts, eco-friendly vehicle subsidies, electric charging infrastructure, and building sector reforms like banning new oil/coal boilers and promoting energy efficiency and sobriety.

33 “Arrêté du 8 décembre 2022 établissant le plan national de réduction des émissions de polluants atmosphériques.” Journal Officiel de la République Française, Texte 27 sur 110.

34 Neither the European Commission nor the European Centre for Medium-Range Weather Forecasts (ECMWF) bears responsibility for the use of Copernicus data in this report.

35 For instance, the point estimate 0.007 for outcome 4 in column (2) in Table 3 when multiplied by 1*/*0.12 equals to 0.058, which is statistically close to 0.057, the corresponding estimate in the fifth row and first column of Table B3, and equates 0.029 when multiplied by 1/0.24 whereas the corresponding estimate in B3 is 0.024.

## Notes

### Competing Interest Statement

The authors have declared no competing interest.

### Funding Statement

- Agence Nationale de la Recherche through the program Investissements d'Avenir ANR-17-EURE-0001
- Social Sciences and Humanities Research Council of Canada

### Author Declarations

The data collection and transmissions for the EDP-Sante to the DREES (Statistical Body of the French Ministry of Solidarity and Health) were approved by the French Data Protection Authority (CNIL) for the project of evaluation of the French National Health Strategy (Strategie Nationale de Sante), which comprises answers to environmental hazards such as air pollution. The DREES is allowed to process personal health data in order to compute statistics, under article 65 of the law Data processing and Liberties (Informatique et Libertes) of January 6th, 1978. All data processing and storage comply with the General Data Protection Regulation (GDPR).

